# Reactive vaccination of workplaces and schools against COVID-19

**DOI:** 10.1101/2021.07.26.21261133

**Authors:** Benjamin Faucher, Rania Assab, Jonathan Roux, Daniel Levy-Bruhl, Cécile Tran Kiem, Simon Cauchemez, Laura Zanetti, Vittoria Colizza, Pierre-Yves Boëlle, Chiara Poletto

**Affiliations:** Sorbonne Université, INSERM, Institut Pierre Louis d’Epidémiologie et de Santé Publique, F75012, France; Univ Rennes, EHESP, REPERES « Recherche en Pharmaco-Epidémiologie et Recours aux Soins » – EA 7449, 15 avenue du Professeur-Léon-Bernard, CS 74312, 35043 Rennes, France; Santé Publique France, Saint Maurice, France; Mathematical Modelling of Infectious Diseases Unit, Institut Pasteur, Université de Paris, UMR2000, CNRS, Paris, France; Collège Doctoral, Sorbonne Université, Paris, France; Haute Autorité de Santé, Saint-Denis, France

## Abstract

As vaccination against COVID-19 stalls in some countries, increased accessibility and more adaptive approaches may be useful to keep the epidemic under control. Here, we study the impact of reactive vaccination targeting schools and workplaces where cases are detected, with an agent-based model accounting for COVID-19 natural history, vaccine characteristics, individuals’ demography and behaviour and social distancing. At an equal number of doses reactive vaccination produces a higher reduction in cases compared with non-reactive strategies, in the majority of scenarios. However, at high initial vaccination coverage or low incidence, few people are found to vaccinate around cases, thus the reactive strategy may be less effective than non-reactive strategies with moderate/high vaccination pace. In case of flare-ups, reactive vaccination could hinder spread if it is implemented quickly, is supported by enhanced test-trace-isolate and triggers an increased vaccine uptake. These results provide key information to plan an adaptive vaccination deployment.

## Introduction

Vaccination against SARS-CoV-2 has changed the course of the COVID-19 pandemic thanks to the high effectiveness of available vaccines in preventing infection and severe forms of the disease. Still, several months into the vaccination campaign vaccine uptake remains below official targets in many Western countries due to logistical issues, vaccine accessibility and/or hesitancy. As of Fall 2021, less than 60% of the population in the United States and Europe was fully vaccinated^1^. With intense virus circulation still ongoing in many regions of the world due to the Delta variant and the threat posed by emerging variants, it is important to investigate if vaccine use could improve with adaptive delivery. Indeed, proposing vaccination to individuals who were exposed to the virus allows targeting those at higher risk of infection and, furthermore, might help overcome barriers to vaccination ^2, 3^ since vaccine-hesitant people are more likely to accept vaccination when the perceived risk of infection is higher^4^.

Redirecting vaccine supplies to geographic areas of highest incidence (or hotspot vaccination) is already part of some European countries’ plans and was implemented to combat the emergence of variant Delta ^2^. But other reactive vaccination schemes are possible, such as ring vaccination that targets contacts of confirmed cases or contacts of those contacts, or vaccination in workplaces or schools where cases have been detected. This could potentially improve vaccine impact by preventing transmission where it is active and even enable the efficient management of flare-ups. For outbreaks of smallpox or Ebola fever, ring vaccination has proved effective to rapidly curtail the spread of cases ^5–8^. However, the experience of these past epidemics cannot be transposed directly to COVID-19 due to the many differences in the infection characteristics and epidemiological context. For example, COVID-19 cases are infectious a few days before symptom onset ^9^, but generally detected a few days later. This means they may have had time to infect their direct contacts, preventing the effectiveness of ring vaccination to prevent secondary cases. Vaccinating an extended network of contacts, as could be done with the vaccination of whole workplaces or schools, would have a larger impact, especially if adopted in combination with strengthened protective measures to slow down transmission, such as masks, physical distancing, and contact tracing. This could be feasible in many countries, leveraging the established test-trace-isolate (TTI) system that enables prompt detection of clusters of cases to inform where vaccines should be deployed. Properly assessing the interest of reactive vaccination therefore requires to consider in detail the interactions of vaccine characteristics, pace of vaccination, COVID-19 natural history, case detection practices and overall changes in human contact behaviour.

We therefore extend an agent-based model that has been previously described ^10^ to quantify the impact of a reactive vaccination strategy targeting workplaces, universities and 12+ years old in schools where cases have been detected. We compare the impact of reactive vaccination with non-reactive vaccination targeting similar settings or with mass vaccination, and test these strategies alone and in combination. We explore differences in vaccine availability and logistic constraints, and assess the influence of the dynamic of the epidemic and different stages of the vaccination campaign.

## Results

### Mass vaccination, targeted, and reactive vaccination strategies

We extend a previously described SARS-CoV-2 transmission model ^10^ to simulate vaccine administration, running in parallel with other interventions - i.e. contact tracing, teleworking and social restrictions. Following similar approaches ^11–14^, the model is stochastic and individual based. It takes as input a synthetic population reproducing demographic, social- contact information, workplace sizes and school types (Figure 1A) of a typical medium-sized French town (117,492 inhabitants). Contacts are described as a dynamic multi-layer network ^10^ (Figure 1B).

**Figure 1.**
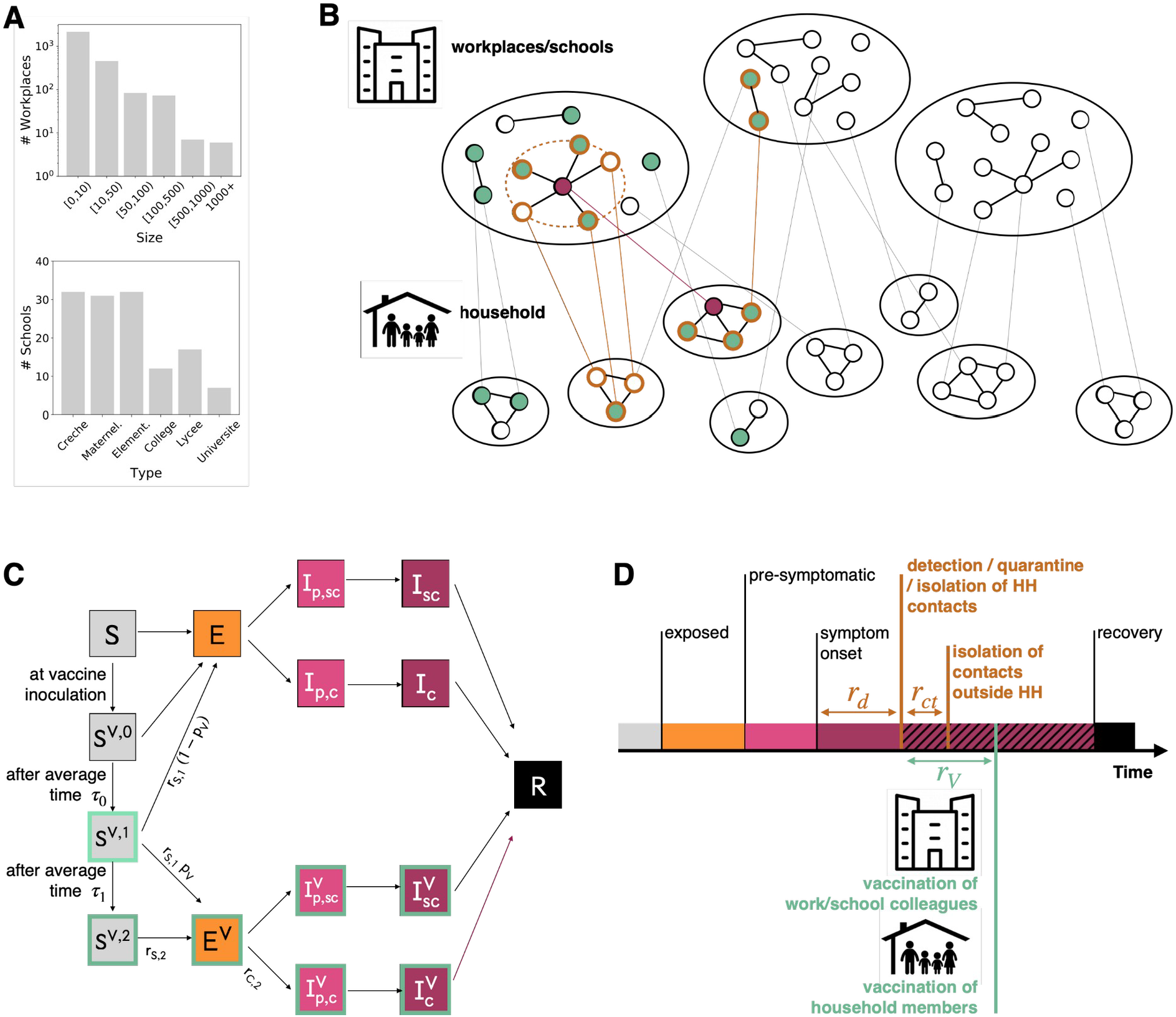
A. Distribution of workplace size and of school type for the municipality of Metz (Grand Est region, France), used in the simulation study. **B** Schematic representation of the population structure, the reactive vaccination and contact tracing. The synthetic population is represented as a dynamic multi-layer network, where layers encode contacts in household, workplace, school, community and transport. In the figure, school and workplace layers are collapsed and community and transport are not displayed for the sake of visualisation. Nodes repeatedly appear on both the household and the workplace/school layer. The identification of an infectious individual (in purple in the figure) triggers the detection and isolation of his/her contacts (nodes with orange border) and the vaccination of individuals attending the same workplace/school and belonging to the same household who accept to be vaccinated (green). **C** Compartmental model of COVID-19 transmission and vaccination. Description of the compartments is reported on the Methods section. **D** Timeline of events following infection for a case that is detected in a scenario with reactive vaccination. For panels C, D transition rate parameters and their values are described in the Methods and in the Supplementary Information.

We assume that the vaccine reduced susceptibility, quantified by the vaccine effectiveness *V E_S_*, and symptomatic illness after infection, quantified by *V E_SP_* ^15^ (Figure 1C). We consider a vaccination strategy based on the Cominarty vaccine ^16^ which is very suitable for reactive vaccination as it is effective, requires only three weeks between the two doses conferring protection and is available in large quantities. We describe the vaccine-induced protection accounting for the vaccine characteristics against the Delta variant - i.e. the dominant variant at the time of writing. Real-life estimates differ, reflecting the complex interplay among the timing of Delta introduction in the population, the co-circulation of other variants, waning of immunity and differential impact by age. In the baseline analysis we consider vaccine effectiveness levels in the middle of the range of estimates provided in a systematic review ^17^. We used a three-week interval between doses as in the vaccine trial. ^16^ For vaccine protection, we conservatively assume that there was no protection in the 2 weeks after the first inoculation, followed by intermediate protection until 2 weeks after the second dose (*V E_S_*_,1_=48 % and *V E_SP_*_, 1_=53 %, see additional details in Table S2 of the Supplementary Information) and maximum protection from this date on (*V E_S_*_,2_=70 % and *V E_SP_*_, 2_=73 %), 5 weeks after the first dose. The maximum protection values are close to the estimates obtained in a meta-analysis for Delta, all vaccines combined ^18^. Lower and higher vaccine effectiveness are also explored.

To parametrise the epidemiological context we assume 32% ^19, 20^ of the population was fully immune to the virus due to previous infection and the reproductive ratio is *R*=1.6, in the range of values estimated for the Delta wave of summer 2021 ^1, 21^. We model the baseline TTI policy after the French situation, allowing 3.6 days on average from symptoms onset to detection and 2.8 average contacts detected and isolated per index case ^22^ (Figure 1 D). We assume that 50% of clinical cases and 10% of subclinical cases are detected, leading to an overall detection rate of ∼25% ^20, 23^. Scenarios with enhanced TTI are described later in the text.

We then model vaccination targeting all adults older than 12 years old with baseline vaccine uptake - set to 80% in the 12-65 years old and 90% in the over 65 years old. ^24^ We assumed that priority risk groups (e.g. elderlies) had already been vaccinated up to that level at the start ^25–28^. We model three *non-reactive* vaccination strategies in the general population, where vaccination is deployed i) randomly throughout the mass vaccination program (*mass*) or ii) in schools sites (*school location,* described below) or iii) in workplaces/universities (*workplaces/universities*) chosen at random, up to the maximum number of doses available daily. In the *school location* vaccination, we assume vaccine sites are created in relation with schools to vaccinate pupils and their household members who are above 12 years old. Then, we model a *reactive vaccination* strategy, where the detection of a case thanks to TTI triggers the vaccination of household members and those in the same workplace or school (Figure 1D). In this scenario, a delay of 2 days on average is assumed between the detection of the case and starting vaccination to account for logistical issues - i.e. ∼5.6 days on average from the index case’s symptoms onset. In the context of reactive vaccination, we test scenarios with vaccine uptake set to its baseline values, and as high as 100%. The impact of each strategy is evaluated based on the comparison with a reference scenario, where no vaccination campaign is conducted during the course of the simulation and vaccination coverage remains at its initial level.

In Figure 2 A, B, C we compare all strategies, assuming that vaccine uptake is the same in reactive and non-reactive vaccination as reference, and initial vaccination coverage is *low*, i.e. ∼30% over the population - with 15% of the [12,60] group and 90% of the 60+ group already vaccinated at the beginning. Similar low coverage values were registered in some countries in Eastern Europe and US counties during the Fall 2021 ^1, 29^. For non-reactive strategies, the vaccination pace is varied between 100 and 500 first doses per 100,000 inhabitants per day. Vaccination pace in Western countries roughly fell within these extremes for the majority of the vaccination campaign, with lower values in general reached around the beginning and the end, due to delivery issues at the beginning, and difficulty in overcoming barriers to vaccination at the end ^1^. For the reactive strategy, vaccine deployment is triggered by detected cases, therefore the number of doses used and the number of places where these doses are administered depends on the epidemic situation. We assume moderate/high initial incidence, i.e. ∼160 clinical cases weekly per 100,000 inhabitants. Panel A shows the relative reduction in the attack rate after two months as a function of the number of first daily doses and Panel B compares the incidence profiles under different strategies with the same number of vaccine doses. The mass, school location and workplaces/universities strategies have a similar impact on the epidemic. They lead to a reduction between 2.7% and 3% of the attack rate with 100 first doses per 100,000 inhabitants administered each day, and between 13% and 15% with 500 per 100,000 inhabitants. The reactive vaccination produces a stronger reduction in cases than the three other strategies in the 2-months period (black dots in Figure 2 A and black line in Figure 2 B). We find that 417 doses per 100,000 inhabitants each day on average are used under the epidemic scenario considered here, yielding an attack-rate relative reduction of 20%.

**Figure 2.**
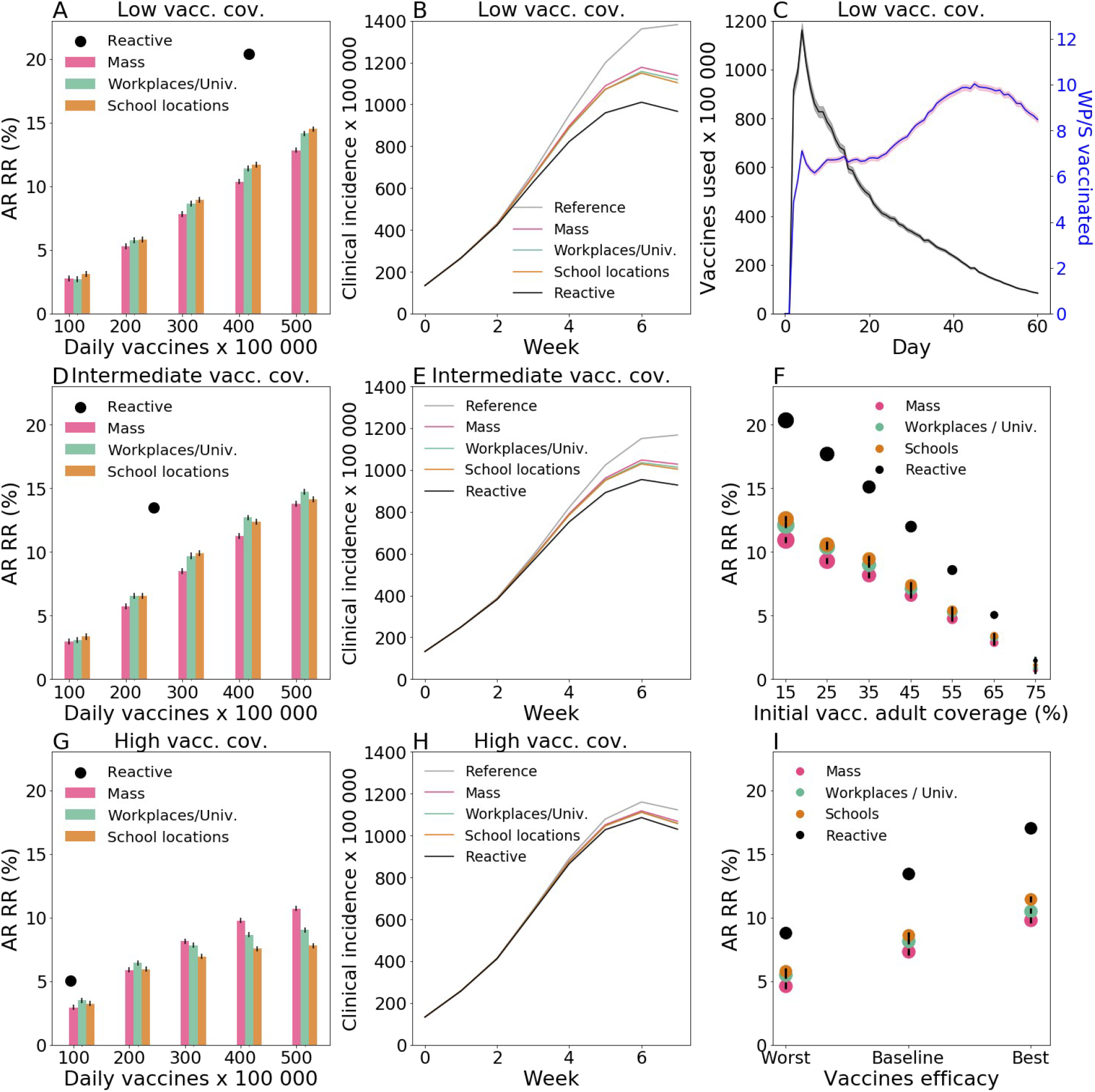
**A, D, G** Relative reduction (RR) in the attack rate (AR) over the first two months for all strategies as a function of the vaccination pace. RR is computed as (ARref - AR)/ARref where ARref is the AR of the reference scenario, where no vaccination campaign is conducted during the course of the simulation and vaccination coverage remains at its initial level. AR is computed from clinical cases. Three initial vaccination coverage are investigated for adults: 15%, i.e. *low vacc. cov.* (A); 40%, i.e. *intermediate vacc. cov.* (D) and 65%, i.e. *high vacc. cov.* (G). **B, E, H** Incidence of clinical cases with different vaccination strategies. The non-reactive scenarios plotted are obtained with the same daily vaccination pace than for reactive vaccination. Low, intermediate and high vaccination coverages are investigated for adults in panels B, E, H, respectively. **C** Number of daily first-dose vaccinations, and number of workplaces/schools (WP/S in the plot) where vaccines are deployed for the same reactive scenario as in A, B - *low vacc. cov.*, with 15% initial vaccine coverage. For the other vaccination coverages the curves are qualitatively similar (results not shown). **F** AR RR for different initial vaccination coverage of adults. The four strategies are compared at equal numbers of vaccine doses. The size of the points is proportional to the average daily vaccination pace. **I** : AR RR for different Vaccine effectiveness levels (VE). The baseline VE values used in the other panels is compared with a worst and a best scenario. The worst scenario is defined by *V E_S_*_,1_=30 %, *V E_SP_*_, 1_=35 %,*V E_S_*_,2_=53 % and *V E_SP_*_, 2_=60 %, while the best scenario is defined by *V E_S_*_,1_=65 %, *V E_SP_*_, 1_=75 %,*V E_S_*_,2_=80 % and *V E_SP_*_, 2_=95 %. For each VE level the four strategies are compared at equal number of vaccine doses. In panel I we consider the *intermediate* vaccination coverage - i.e. 40% initial coverage among adults. In all panels we assumed the following parameters: R=1.6; Initial immunity 32%; vaccinated at the beginning 90% for 60+; 10% of individuals are doing teleworking and 5% of community contacts are removed. Values are means over 2000 stochastic simulations. Error bars in panels A, D, G, F, I are derived from the standard errors of the AR. These are smaller than the size of the dots in almost all cases. Shaded areas in panels B, C, E, H indicate the standard errors - very low in panels B, E, H.

In panel C we consider the same parametrization as in Figure A, B and we show the number of first doses in time and the number of places to vaccinate - as a proxy to the incurred logistics of vaccine deployment. The number of daily inoculated doses is initially high, with almost 1200 doses per 100,000 inhabitants used in a day at the peak of vaccine demand, but declines rapidly afterwards down to 6 doses. The number of workplaces to vaccinate follows a different trend. It slowly increases to reach a peak and then declines. The breakdown in Figure S3 of the Supplementary Information shows that schools and large workplaces are vaccinated at the very beginning. Thus a great number of vaccines are initially deployed in large settings, requiring many doses, while as the epidemic spreads it reaches a large number of small settings where only a few individuals can be vaccinated.

In panels D, E we consider an *intermediate* vaccination coverage at the beginning (40% of the [12,60] group and 90% of 60+, corresponding to ∼45% of the whole population). Non-reactive strategies lead to a relative reduction in the attack rate after two months close to the low initial coverage case. Instead, the impact of reactive vaccination is reduced. A smaller proportion of people attending workplaces/schools that are targeted by vaccination are left to vaccinate, therefore less vaccines are deployed with reduced impact on the epidemic at the population level (Figure 2 D). Still, with ∼250 doses per 100,000 inhabitants each day used on average, reactive vaccination produced a 13% reduction in the attack rate. This value is close to the reduction level reached with non-reactive strategies with 400 doses per 100,000 inhabitants each day. The impact of reactive vaccination becomes very small when initial vaccination coverage is *high*. Panels G, H show a scenario where 65% of the [12,60] group and 90% of 60+ is vaccinated at the beginning, corresponding to ∼60% of the whole population close to the coverage reached in Europe in the Fall 2021 ^1^. Only 94 daily vaccines per 100000 inhabitants are deployed each day with a 5% reduction in the attack rate compared to the 2% reduction of non-reactive strategies at equal number of doses. Non-reactive strategies with vaccination pace higher or equal to 300 doses per 100,000 inhabitants each day produce a higher reduction in cases (∼8% or higher).

The effect of the initial vaccination coverage on the impact of the different strategies is summarized in Figure 2 F. The relative reduction declines roughly linearly with the initial vaccination coverage. The reactive vaccination always outperforms non-reactive strategies at equal number of doses. Still, the number of vaccinated people in the reactive vaccination progressively decreases as initial vaccination coverage increases, to reach the point in which the strategy is less effective compared to non-reactive strategies with not-small vaccination pace. Eventually, in panel I we explore different levels of vaccine effectiveness encompassing the whole range of estimates provided by real life estimates ^17^. We find that lower vaccine effectiveness values lead to a reduced effect of vaccination as expected. The difference between reactive and non-reactive strategies is also reduced.

In the Supplementary Information we analyse the comparison between reactive and non-reactive strategies with varying key parameters. In Figure S4 we assumed the baseline scenario of Figure 2 D, E, i.e. intermediate vaccination coverage at the beginning - ∼45% of the whole population vaccinated. We explore alternative values of transmission, incubation period, immunity level of the population, reduction in contacts due to social distancing, time needed for the vaccine to become effective, compliance to vaccination and vaccine effect on the infection duration. Increase in the reproductive ratio, initial immunity and time between doses reduce the impact of the reactive vaccination. An increase in compliance to vaccination, instead, enhanced the impact of both reactive and non-reactive vaccination.

Other parameters had a more limited role on strategies’ effectiveness. We then consider a scenario of a flare-up of cases, as it may be caused by a new variant of concern (VOC) spreading in the territory (Figure S5). The deployment of vaccines is in this case limited and slow. Depending on the values of the other parameters, i.e. proportion of teleworking and time from building immunity following vaccination, the reactive strategy may bring limited or no benefit with respect to non-reactive strategies, when the comparison is done at equal number of doses. Eventually, we test the robustness of our results according to the selected health outcome, using hospitalisations, ICU admissions, ICU bed occupancy, deaths, life-years lost and quality-adjusted life-years lost, finding the same qualitative behavior.

### Combined reactive and mass vaccination for managing sustained COVID-19 spread

With high availability of vaccine doses, reactive vaccination could be deployed on top of mass vaccination. We consider the *intermediate* vaccination coverage scenario (i.e. ∼45% of the whole population vaccinated) defined in the previous section and compare mass and reactive vaccination simultaneously (*combined strategy*) with mass vaccination alone. We focus on the first two months since the implementation of the vaccination strategy. At an equal number of doses within the period, the combined strategy outperforms mass vaccination in reducing the attack rate. For instance, the relative reduction in the attack rate would go from 10%, when ∼360 daily doses per 100,000 inhabitants are deployed for mass vaccination, to 16%, when the same number of doses are used for reactive and mass vaccination combined (Figure 3A).

**Figure 3.**
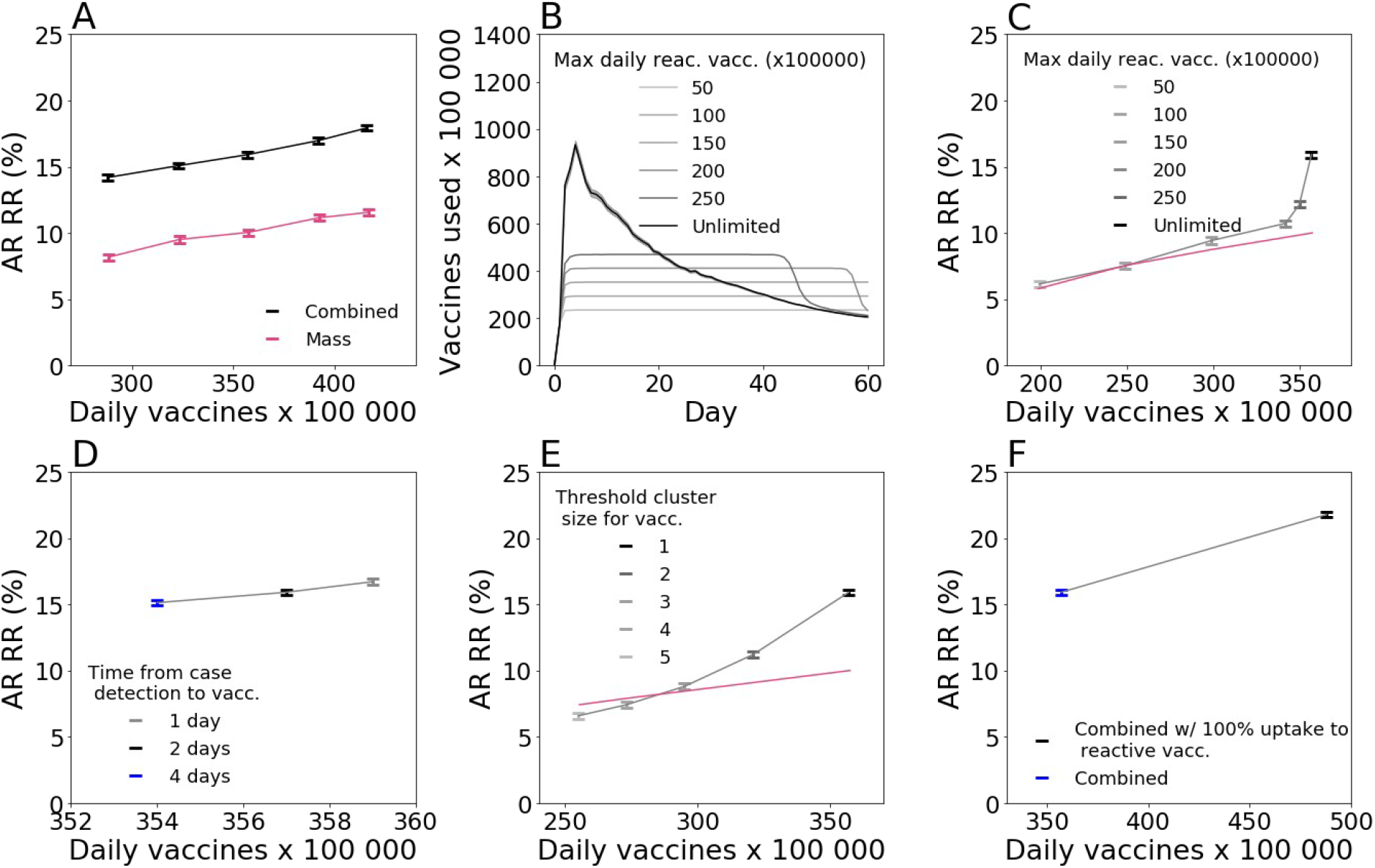
**A** Relative reduction (RR) in the attack rate (AR) over the first two months for the combined strategy (mass / reactive) and the mass strategy with the same number of first-dose vaccinations as in the combined strategy during the period. RR is computed with respect to the reference scenario with initial vaccination only, as in Figure 2. Combined strategy is obtained by running in parallel the mass strategy - from 50 to 250 daily vaccination rate per 100,000 inhabitants - and the reactive strategy. Number of doses displayed in the x-axis of the figure is the total number of doses used by the combined strategy daily. Corresponding incidence curves are reported in Figure S7 of the Supplementary Information. **B** Number of first-dose vaccinations deployed each day for the combined strategy with different daily vaccines’ capacity limits. **C, D, E** AR RR for the combined strategy as a function of the average daily number of first-dose vaccinations in the two-month period. Symbols of different colours indicate: (**C**) different values of daily vaccines’ capacity limit; (**D**) different time from case detection to vaccine deployment; (**E**) different threshold size for the cluster to trigger vaccination. In panel C and E the curve corresponding to mass vaccination only is also plotted for comparison. **F** Comparison between 100% and baseline vaccination uptake in case of reactive vaccination. In all cases we assumed the following parameters: R=1.6; Initial immunity 32%; vaccinated at the beginning 90% and 40% for 60+ and <60, respectively; 10% of individuals are doing teleworking and 5% of community contacts are removed. Values are means over 2000 stochastic simulations, error bars are derived from the standard errors of the AR. In panel B only the standard error of the unlimited case is shown, this indicated by the shaded area.

We explore alternative scenarios where the number of vaccines used and places vaccinated are limited due to availability and logistic constraints. We assess the effect of three parameters: (i) the maximum daily number of vaccines that can be allocated towards reactive vaccination (with caps going from 50 to 250 per 100,000 inhabitants, compared with unlimited vaccine availability assumed in the baseline scenario), (ii) the time from the detection of a case and the vaccine deployment (set to 2 days in the baseline scenario, and here explored between 1 and 4 days), and (iii) the number of detected cases that triggers vaccination in a place (from 2 to 5 cases, vs. the baseline value of 1).The number of first-dose vaccinations in time under the different caps is plotted in Figure 3 B. A small cap on the number of doses limits the impact of the reactive strategy. Figure 3 C shows that the attack rate relative reduction drops from 16% to 6% if only a maximum of 50 first doses per 100,000 inhabitants daily can be used in reactive vaccination, reaching the levels of mass vaccination only.

Doubling the time required to start reactive vaccination, from 2 days to 4 days, has a limited effect on the reduction of the AR (relative reduction reduced from 16% to 15%, Figure 3D). Increasing the number of detected cases used to trigger vaccination to 2 (respectively 5) reduces the relative reduction to 11% (respectively 6%) (Figure 3E).

We so far assumed that vaccine uptake is the same in mass and reactive vaccination. This assumption is likely conservative, in that individuals may be more inclined to accept vaccination when this is proposed in the context of reactive vaccination due to the higher perceived benefit of vaccination. In Figure 3F we consider a scenario where vaccine uptake with reactive vaccination climbs to 100%. Attack rate relative reduction increases in this case from 16% to 22%, with a demand of ∼480 daily doses per 100,000 inhabitants on average.

### Combined reactive and mass vaccination for managing a COVID-19 flare-up

We have previously said that in a scenario of flare-up of cases reactive vaccination would bring limited benefit compared to other strategies (Figure S5 of the Supplementary Information). Here we analyse this scenario more in depth, assuming that reactive vaccination is combined with mass vaccination, but triggers an increase in vaccine uptake and is associated with enhanced TTI, as may be the case in a realistic scenario of alert due to initial VOC detection.

We assume mass vaccination with 150 first doses per day per 100,000 inhabitants is underway from the start, with *intermediate* vaccination coverage at the beginning (i.e. ∼45% of the whole population vaccinated). We assume baseline TTI is in place prior to cases’ introduction. To start a simulation, three infectious individuals carrying a VOC are introduced in the population where the virus variant is not currently circulating. Upon detection of the first case, we assume that TTI is enhanced, finding 70% of clinical cases, 30% of subclinical cases (i.e. ∼45% of all cases) and three times more contacts outside the household with 100% compliance to isolation (Table S4 of the Supplementary Information) - the scenario without TTI enhancement is also explored for comparison. As soon as the number of detected cases reaches a predefined threshold, reactive vaccination is started on top of the mass vaccination campaign. We assume vaccine uptake increases to 100% for reactive vaccination but stays at its baseline value for other approaches.

In Figure 4 we compare the combined scenario with mass vaccination alone at an equal number of doses, and we investigate starting the reactive vaccination after 1, 5 or 10 detected cases. Panels A, B show the case of baseline transmissibility and vaccine effectiveness. With reactive vaccination starting from the first detected case, the attack rate decreases by ∼10%, compared with the mass scenario. However, the added value of reactive vaccination decreases if more detected cases are required to start the intervention. Without enhancement in TTI and increase in vaccine uptake, attack rate values are much higher and the benefit of reactive vaccination over the mass vaccination is lower (∼3%).

**Figure 4.**
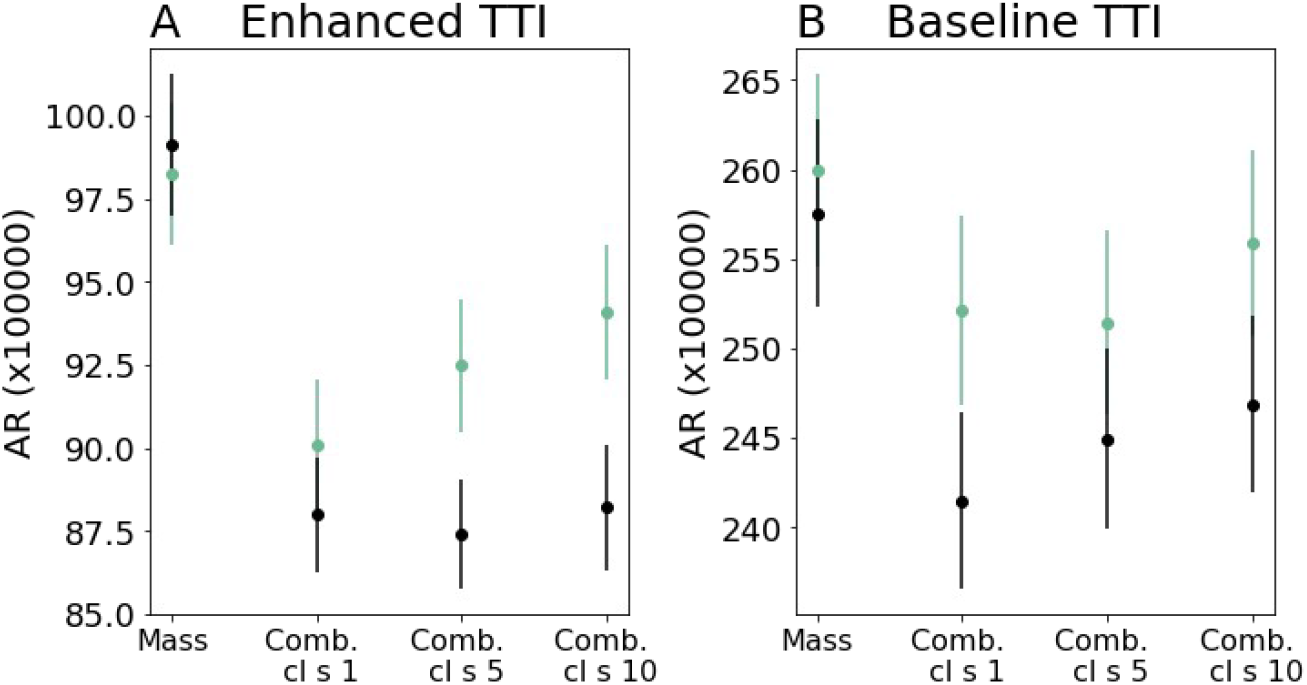
**A, B** Average attack rate per 100000 inhabitants in the first two months for the enhanced (A) and baseline (B) TTI scenario. Four vaccination strategies are compared: mass only, combined where the reactive vaccination starts at the detection of 1, 5, 10 cases (Comb. cl. s.= 1, 5, 10 in the figure). For mass vaccination the number of first-dose vaccinations during the period is the same as in the comb. cl. s=1 of the same scenario. In all panels we assume the following parameters: R=1.6; Initial immunity 32%; 90% for 60+ year old and 40% for <60 years old already vaccinated at the beginning; 10% of individuals are doing teleworking and 5% of community contacts are removed. Corresponding incidence curves are reported in Figure S8 of the Supplementary Information. Error bars are the standard errors from 8000 stochastic realisations.

In the Supplementary Information we consider different epidemic scenarios, testing different values for the transmissibility and vaccine effectiveness - including worst-case vaccine effectiveness, and R as high as 1.8 -, finding similar trends. Eventually, we analysed the impact of vaccination on the flare-up extinction. With the parameterization of Figure 4 A we find probabilities of extinction equal to ∼5.5% and ∼6% with mass and combined strategies, respectively. These values respectively become ∼15% and ∼18% in a best-case scenario with a more rapid mounting of vaccine effect after the first dose, best-case vaccine effectiveness and strong TTI.

## Discussion

The rapid rise of new SARS-CoV-2 variants with increased transmissibility has made the course of the COVID-19 pandemic unpredictable, posing a persistent public health threat that jeopardises relaxation of NPIs and return to normal life ^25, 30–34^. More transmissible viruses call for vaccination of a larger proportion of the population, therefore vaccination must be made more accessible and able to adapt to a rapidly evolving epidemic situation ^2^. In this context, we have analysed the reactive vaccination of workplaces, universities and schools to assess its potential role in managing the epidemic.

We presented an agent-based model that accounts for the key factors affecting the effectiveness of reactive vaccination: disease natural history, vaccine characteristic, individual contact behaviour, and logistic constraints. For a wide range of epidemic scenarios, the reactive vaccination had a stronger impact on the COVID-19 epidemic compared to non-reactive vaccination strategies (including the standard mass vaccination) at equal number of doses used within the two months after inception. In addition, combining reactive and mass vaccination was more effective than mass vaccination alone. For instance, in a scenario of moderate/high incidence with ∼45% vaccination coverage at the beginning we found that the relative reduction in the attack rate after two months would improve from 10% to 16% with ∼350 daily first vaccine doses per 100000 habitants used in a combined mass/reactive vaccination approach instead of mass only. However, reactive vaccination had limited or no advantage with respect to non-reactive strategies under certain circumstances, as the number of doses administered with the reactive vaccination depended on the number and pace of occurrence and detection of COVID-19 cases. This may be the case when vaccination coverage is already high at the beginning and only a few people to vaccinate are found around detected cases, or in a flare-up scenario when only a few cases are detected. Non-reactive strategies could then be more effective as long as the pace of vaccine administration is not small. Yet, in these situations, adding reactive vaccination to mass vaccination could become of interest again by triggering an increase in vaccine uptake, all the more if this is combined with enhanced TTI.

Reactive vaccination has been studied for smallpox, cholera and measles, among others ^5–7, 35, 36^. Hotspot vaccination was found to help in cholera outbreak response by both modelling studies and outbreak investigation ^36, 37^. It may target geographic areas defined at spatial resolution as diverse as districts within a country, or neighbourhoods within a city, according to the situation. For Ebola and smallpox ring vaccination was successfully adopted to accelerate epidemic containment ^5–7^. For these infections, vaccine-induced immunity mounts rapidly compared to the incubation period and contacts of an index case can be found before they start transmitting since pre-symptomatic and asymptomatic transmission is almost absent. Ring vaccination is also likely effective when the vaccine has post-exposure effects^8^. Reactive vaccination of schools and university campuses has been implemented in the past to contain outbreaks of meningitis ^38^ and measles ^39, 40^.

For COVID-19, the use of reactive vaccination has been reported in Ontario, the UK, Germany, France, among others ^2, 41–46^. In these places, vaccines were directed to communities, neighbourhoods or building complexes with a large number of infections or presenting epidemic clusters or surge of cases due to virus variants. The goal of these campaigns was to minimise the spread of the virus, but it also addressed inequalities in access and increased fairness, since a surge of cases may happen where people have difficulty in isolating due to poverty and house crowding ^47^. In France, reactive vaccination was implemented to contain the emergence of variants of concerns in the municipalities of Bordeaux, Strasbourg and Brest ^44–46^. In the municipality of Strasbourg, vaccination slots dedicated to students were created following the identification of a Delta cluster in an art school ^44^. Despite the interest in the strategy and its inclusion in the COVID-19 response plans, very limited work has been done so far to quantify its effectiveness ^48, 49^. A modelling study on ring-vaccination suggested that the strategy could be valuable if the vaccine has post-exposure efficacy and a large proportion of contacts could be identified ^49^. Still, post-exposure effects of the vaccine remain currently under-investigated, ^50^ and it is likely that the vaccination of the first ring of contacts alone would bring little benefit, if at all. We have here tested reactive vaccination of workplaces and schools, since focusing on these settings may be an efficient way to easily reach an extended group of contacts. Workplaces have been found to be an important setting for COVID-19 transmission, especially specific workplaces where conditions are more favourable for spreading ^51, 52^. University settings also cover a central role in the COVID-19 transmission, due to the higher number of contacts among students, particularly if sharing common spaces in residence accommodations ^53^. Model results show that reactive vaccination of these settings could have in many circumstances a stronger impact than simply reinforcing vaccination as in hotspot strategies.

However, the effectiveness of the reactive strategy depends on the epidemic context. We found that, as vaccination coverage increases, the relative advantage of reactive vaccination over non-reactive strategie diminishes. For >40% vaccination coverage among adults, the relative reduction of the strategy is smaller than the one produced by non-reactive vaccination at a moderate/high vaccination pace, under the hypothesis of no increase in vaccine uptake in the context of reactive vaccination. As discussed before, when a cluster is identified in a given workplace/school the vaccine is proposed only to a few individuals not previously vaccinated and compliant to vaccination, resulting in a limited deployment of vaccine and a reduced impact. In addition, breakthrough infection becomes an important driver of propagation when a large proportion of individuals are vaccinated, with consequently a larger proportion of subclinical cases and in turn a reduced detection rate overall. This makes the detection of outbreaks and consequently the reactive deployment of vaccines more difficult.

If initial vaccination coverage is not too high, the feasibility and advantage of the inclusion of reactive vaccination imply a trade-off between epidemic intensity and logistic constraints. At a moderate/high incidence level, combining reactive and mass vaccination would substantially decrease the attack rate compared to mass vaccination for the same number of doses, but the large initial demand in vaccines may exceed the available stockpiles. Even with large enough stockpiles, issues like the timely deployment of additional personnel in mobile vaccine units and the need to quickly inform the population by communication campaigns must be solved to guarantee the success of the campaign. We explored with the model the key variables that would impact the strategy effectiveness. Delaying the deployment of vaccines in workplaces/schools upon the detection of a case (from 2 to 4 days on average) would not have a strong impact on its effectiveness. However, vaccines should be deployed at the detection of the first case to avoid substantially limiting the impact of the strategy - e.g. the relative reduction goes from 16% to 6% when workplaces/schools are vaccinated at the detection of 5 cases (Figure 3E).

In the case of a COVID-19 flare-up the reactive strategy may bring an advantage if the reactive strategy starts early, it is combined with increased TTI and it triggers an increase in vaccine uptake. First, an early start of the reactive vaccination campaign since the detection of the first VOC case requires that tests for the detection of variants must be carried out quickly and with large coverage. Genomic surveillance has ramped up in many countries since the emergence of the Alpha variant in late 2020. In France, nationwide surveys are conducted every two weeks involving the full genome sequencing of randomly selected positive samples ^54^. Approximately 50% of positive tests are also routinely screened for key mutations to monitor the circulation of the main variants registered as VOC or VUI ^54^. While these volumes of screening may be regarded as sufficient to quickly identify the presence of variants, the quick rising of large clusters of cases is possible, notably due to super spreading events, becoming increasingly frequent as social restrictions relax. Second, reactive vaccination must be part of a wider response plan, including notably a strong intensification of TTI ^23^. Rapid and efficient TTI efficiently mitigates spread on its own, but it is also instrumental to the success of reactive vaccination as it allows triggering vaccination in households, workplaces and schools. Third, an increased level of vaccine uptake is essential for reactive vaccination to be of interest. In many Western countries, upward trends in vaccine uptake have been observed as the vaccination campaign unfolds, thanks to government mandates, incentives by public health authorities, the communication effort and the strong evidence regarding vaccine efficacy. Still vaccination coverage is highly heterogeneous and remains low, e.g., in many countries of Eastern Europe and in many counties in the US ^1, 29^. Besides the individuals who oppose vaccination, a reactive strategy may have the potential to increase acceptability of the vaccine by making it more accessible and anticipating an immediate benefit against the risk of infection. An increase in vaccine uptake was indeed observed in the context of a reactive vaccination campaign during the course of a measles outbreak ^4^.

Therefore reactive vaccination could be an important way to improve access to vaccination - especially for the hard-to-reach population - and potentially increase acceptability, e.g. due to risk perception.

The study is affected by several limitations. First, the synthetic population used in the study accounts for the repartition of contacts across workplaces, schools, households, etc., informed by contact surveys. However, numbers of contacts and risk of transmission could vary greatly according to the kind of occupation. The synthetic population accounts for this variability assuming that the average number of contacts from one workplace to another is gamma distributed ^10^. Still, no data were available to inform the model in this respect. Second, we model vaccination uptake according to age only, when it is determined by several sociodemographic factors. Clusters of vaccine hesitant individuals may play an important role in the dynamics and facilitate the epidemic persistence in the population, as it is described for measles ^55^. In those countries where vaccination coverage is high, heterogeneities in attitude toward vaccination may have an impact. Third, the agent-based model is calibrated from French socio-demographic data. The results of this study can be extended to countries with similar societal structure and contact patterns, as e.g. other developed countries ^56^. Still, COVID-19 transmission potential, level of disease-induced immunity, vaccination coverage, and extent of social restrictions vary substantially from one country to another. In addition, the time since vaccination campaign was started, the consequent extent of waning of immunity and countries’ strategies of administering the vaccine booster affect the level of protection of the population already vaccinated and may alter the impact of reactive deployment of vaccines among individuals not yet vaccinated. We explored a large set of parameters in the Supplementary Information to provide fundamental understanding on the interplay between the epidemic and the reactive vaccination dynamics that may aid the planning of the strategy in case of future epidemic surges.

## Methods

### Synthetic population

We use a synthetic population for a French municipality based on the National Institute of Statistics and Economic Studies (INSEE) censuses and French contact survey information ^10, 57^. This includes the following input files: i) a setting-specific, time-varying network of daily face-to-face contacts; ii) the maps between individuals and their age, iii) between individuals and the household they belong to, iv) between individuals and their school, v) and between individuals and their workplace. The synthetic population has age pyramid, household composition, number of workplaces by size, and number of schools by type, reproducing INSEE statistics. Daily face-to-face contacts among individuals are labelled according to the setting in which they occur (either household, workplace, school, community or transport) and they have assigned a daily frequency of activation, to explicitly model recurrent and sporadic contacts. We consider the municipality of Metz in the Grand Est region, which has 117,492 inhabitants, 131 schools (from kindergarten to University) and 2888 workplaces (Figure 1A). Detailed description of how the population was generated is provided in ^10^. Information about how to access population files is provided in the Data availability section.

### Overview of the model

The model is written in C/C++, and is stochastic and discrete-time. It accounts for the following components: (i) teleworking and social distancing, (ii) COVID-19 transmission, accounting for the effect of the vaccine; (iii) test-trace-isolate; (iv) vaccine deployment. Model output includes time series of incidence (clinical and subclinical cases), detailed information on infected cases (time of infection, age, vaccination status), vaccines administered according to the strategy, number of workplaces where vaccines are deployed. Different epidemic scenarios are explored and compared. In the Supplementary Information we also analyse hospitalisation entries, deaths, ICU entries, life-year lost, quality-adjusted life-year, ICU bed occupancy. These quantities are computed by postprocessing output files containing the detailed information on infected cases.

### Teleworking and social distancing

Teleworking and other social restrictions may alter the repartition of contacts across settings and in turn the effectiveness of vaccination strategies ^23^. We thus explicitly accounted for this ingredient in the model. Specifically, to model teleworking we assume a proportion of nodes are absent from work, modelled by erasing working contacts and transport contacts of these nodes. To account for the reduction in social encounters due to the closure of restaurants and other leisure activities we removed a proportion of contacts from the community layer. In Western countries, level of restrictions varied greatly both by country and in time since vaccines were first deployed at the beginning of 2021. We set the contact reduction in the community to 5% and the teleworking to 10%. These are close to the reduction values reported by google mobility reports for France during Autumn 2021 ^58^, and fall within the range of European countries’ estimates. Note that levels of teleworking ∼10% for European countries are reported also by other sources ^59^. Scenarios with different levels of contact reduction are compared in the Supplementary Information. Telework and social distancing is implemented at the beginning of the simulation and remains constant for the duration of the simulation. Importantly, the reproductive ratio is set to the desired value, independently by the level of contact reduction, as described in the Supplementary Information.

### COVID-19 transmission model

Transmission model is an extension of the model in ^10^ (see Figure 1D). This accounts for heterogeneous susceptibility and severity across age groups ^60, 61^, the presence of an exposed and a pre-symptomatic stage ^9^, and two different levels of infection outcome - subclinical, corresponding to asymptomatic infection and paucisymptomatic, and clinical, corresponding to moderate to critical infection ^60, 62^. Precisely, susceptible individuals, if in contact with infectious ones, may get infected and enter the exposed compartment (*E*). After an average latency period *_ɛ_* ^−1^they become infectious, developing a subclinical infection (*I _sc_*) with age- dependent probability *p^A^* and a clinical infection (*I _c_*) otherwise. From *E*, before entering either *I _sc_* or *I _c_*, individuals enter first a prodromal phase (either *I _p,sc_* or *I _p,c_*), that lasts on average *μ*^−1^ days. Compared to *I _p,c_* and *I _c_* individuals, individuals in the *I _p,sc_* and *I _sc_* compartments have reduced transmissibility rescaled by a factor *β_I_*. With rate *μ* infected individuals become recovered. Age-dependent susceptibility and age-dependant probability of clinical symptoms are parametrised from ^60^. In addition, transmission depends on setting as in ^10^. We assume that the time spent in the *E*, *I _p,sc_* and *I _p,c_*, is Erlang distributed with shape 2, and rate 2 *ɛ* for *E*, and 2*μ_p_* for *I _p,sc_* and *I _p,c_*. Time spent in *I _sc_* and *I _c_*is exponentially distributed. Parameters are summarised in Table S1 of the Supplementary Information.

We model vaccination with a leaky vaccine, partially reducing both the risk of infection (i.e. reduction in susceptibility, *V E_S_*) and infection-confirmed symptomatic illness (*V E_SP_*) ^15^. Level of protection increases progressively after the inoculation of the first dose. In our model we did not explicitly account for the two-dose administration, but we accounted for two levels of protection – e.g. a first one approximately in between of the two doses and a second one after the second dose. Vaccine efficacy was zero immediately after inoculation, mounting then to an intermediate level (*V E_S_*_,1_ and *V E_SP_*_, 1_) and a maximum level later (*V E_S_*_,2_ and *V E_SP_*_, 2_).

This is represented through the compartmental model in Figure 1C. Upon administering the first dose, *S* individuals become, *_S_^V,^*^0^, i.e. individuals that are vaccinated, but have no vaccine protection. If they do not become infected, they enter the stage *_S_^V,^* ^1^, where they are partially protected, then stage *_S_^V,^* ^2^ where vaccine protection is maximum. Time spent in *_S_^V,^* ^0^and *_S_^V,^* ^1^ is Erlang distributed with shape 2 and rate 2/ *τ* _0_and 2/ *τ*_1_ for *_S_^V,^*^0^ and *_S_^V,^* ^1^, respectively. *_S_^V,^* ^1^ and *S^V,^* ^2^ individuals have reduced probability of getting infected by a factor *r _S_*_,1_=(1−*V E_S_*_,1_) and *r _S_*_,2_=(1−*V E_S_*_,2_), respectively. In case of infection, *_S_^V,^* ^2^ individuals progress first to exposed vaccinated (*E^V^*), then to either preclinical or pre-subclinical vaccinated (*I^V^_p,c_* or *I ^V^_p,sc_*) that are followed by clinical and subclinical vaccinated respectively (*I^V^_c_* or *I ^V^_sc_*). Probability of becoming *I^V^_c_,* 2 from *_E_^V^* is reduced of a factor *r_c_*_, 2_=(1−*V E_SP_*_,2_)(1−*V E_S_*_,2_)^−1^. For the *S_V_*_, 1_ individuals that get infected we assume a polarised vaccine effect, i.e. they can enter either in *_E_^V^*, with probability *p_V_*, or in *E* (Figure 1D). The value of *p_V_* can be set based on *V E_SP_*_, 1_ through the relation (1−*V E_SP_*_,1_)=(1−*V E_S_*_,1_) (*p_V_ r_c_*_,2_ +(1−*p_V_*)).We assume no reduction in infectiousness for vaccinated individuals. However, we accounted for a 25% reduction in the duration of the infectious period as reported in ^63, 64^.

Under the assumption that no serological/virological/antigenic test is done before vaccine administration, the vaccine is administered to all individuals, except for clinical cases who show clear signs of the disease or individuals that were detected as infected by the TTI in place. In our model a vaccine administered to infected or recovered individuals has no effect.

In the baseline scenario we parametrise *V E_SP_*_, 1_, *V E_SP_*_, 2_, *V E_S_*_,1_and *V E_S_*_,2_ by taking values in the middle of estimates reported in the systematic review by Higdon and collaborators ^17^ for the Comirnaty vaccine and the Delta variant ^65^. Chosen values of *V E_SP_*_, 2_, and *V E_S_*_,2_are also comparable with the effectiveness estimates reported in a meta-analysis for the Delta variant, complete vaccination, all vaccines combined ^18^. We also test values on the upper and lower extremes of the range of estimates of ^17^. Parameters are listed in Table S2 of the Supplementary Information.

### Test-trace-isolate

We model a baseline TTI accounting for case detection, household isolation and manual contact tracing. Fifty percent of individuals with clinical symptoms were assumed to get tested and to isolate if positive. We assume an exponentially distributed delay from symptoms onset to case detection and its isolation with 3.6 days on average. Once a case is detected, his/her household members isolate with probability *p_ct,HH_*, while other contacts isolate with probabilities *p_ct,A_* and *p_ct,Oth_*, for acquaintances and sporadic contacts, respectively. In addition to the detection of clinical cases, we assume that a proportion of subclinical cases were also identified (10%). Isolated individuals resume normal daily life after 10 days unless they still have clinical symptoms after the time has passed. They may, however, decide to drop out from isolation each day with a probability of 13% if they do not have symptoms ^66^.

In the scenario of virus re-introduction we consider enhanced TTI, corresponding to a situation of case investigation, screening campaign and sensibilisation (prompting higher compliance to isolation). We assume a higher detection of clinical and subclinical cases (70% and 30%, respectively), perfect compliance to isolation by the index case and household members and a three-fold increase in contacts identified outside the household.

Step-by-step description of contact tracing is provided in the Supplementary Information. Parameters for baseline TTI are provided in Table S3, while parameters for enhanced TTI are provided in Table S4.

### Vaccination strategies

A vaccine opinion (willingness or not to vaccinate) is stochastically assigned to each individual at the beginning of the simulation depending on age (below/above 65 years old). Opinion does not change during the simulation. In some scenarios we assume that all individuals are willing to accept the vaccine in case of reactive vaccination, while maintaining the opinion originally assigned to them when the vaccine is proposed in the context of non- reactive vaccination. Only individuals above a threshold age, *a_th, V_* =12 years old, are vaccinated. We assume that a certain fraction of individuals are vaccinated at the beginning of the simulation according to the age group ([12,60], 60+). We compare the following vaccination strategies:

*Mass*: *V_daily_* randomly selected individuals are vaccinated each day until a *V _tot_* limit is reached.

*Workplaces/universities*: Random workplaces/universities are selected each day. All individuals belonging to the place, willing to be vaccinated, and not isolated at home that day are vaccinated. Individuals in workplaces/universities are vaccinated each day until the daily limit, *V_daily_* is reached. No more than *V _tot_* individuals are vaccinated during the course of the simulation. We assume that only workplaces with ¿¿ *th*=20 ¿ employees or larger implement vaccination.

*School location*: Random schools, other than universities, are selected each day and a vaccination campaign is conducted in the places open to all household members of school students. All household members willing to be vaccinated, above the threshold age and not isolated at home that day are vaccinated. No more than *V _daily_* individuals are vaccinated each day and no more than *V _tot_* individuals are vaccinated during the course of the simulation.

*Reactive*: When a case is detected, vaccination is done in her/his household with rate *r_V_*^❑^. When a cluster – i.e. at least *n_cl_* cases detected within a time window of length *T _cl_* – is detected in a workplace/school, vaccination is done in that place with rate *r_V_*^❑^.. In the baseline scenario, we assume vaccination in workplace/school to be triggered by one single infected individual (*n_cl_*=1). In both household and workplace/school, all individuals belonging to the place above the threshold age and willing to be vaccinated are vaccinated. Individuals that were already detected and isolated at home are not vaccinated. No more than *V_daily_* individuals are vaccinated each day and no more than *V _tot_* individuals are vaccinated during the course of the simulation. In the baseline scenario these quantities are unlimited, i.e. all individuals to be vaccinated in the context of reactive vaccination are vaccinated.

Parameters and their values are summarised in Table S5 of the Supplementary Information.

### Data availability

The synthetic population used in the analysis is available on github ^67^.

### Code availability

We provide all C/C++ code files of the model on github ^67^.

## Data Availability

Code and input files of the study are posted on the github repository

https://github.com/PYB-SU/ReactiveVaccination

## Acknowledgements

We acknowledge financial support from Haute Autorité de Santé; the ANR and Fondation de France through the project NoCOV (00105995); the Municipality of Paris (https://www.paris.fr/) through the programme Emergence(s); EU H2020 grants MOOD (H2020-874850), and RECOVER (H2020-101003589); the ANRS through the project EMERGEN (ANRS0151); the ANR through the project SPHINX (ANR-17-CE36-0008-05); Institut des Sciences du Calcul et de la Donnée.Supplementary Information

## Supplementary Information

### Additional Methods

#### COVID-19 transmission model

We provide here in the following the parameter values for transmission and infection natural history (Table S1), and the effect of vaccination (Table S2). For a detailed explanation of the transmission model without vaccination we refer to ^1^. Incubation period *IP* and length of the pre-symptomatic phase *μ_p_* are specific for the Delta variant. In particular, *μ_p_* is prametrised based on the proportion of pre-symptomatic transmission estimated in ^2^.

**Table S1.** Transmission parameters and their baseline values.

| Parameter | Description | Baseline value (other explored values) | Source |
| --- | --- | --- | --- |
| $IP$ | Incubation period | 5.8 days ( 5.1 days, 6.3 days) | <sup>2</sup> (<sup>3,4</sup>) |
| $\mu_p$ | Rate of developing symptoms for pre-symptomatic individuals | (2.1 days)<sup>-1</sup> | Computed to recover 74% of pre-symptomatic transmission as estimated in <sup>2</sup> |
| $\epsilon$ | Rate of becoming infectious for exposed individuals | (3.7 days)<sup>-1</sup> | $IP - \mu_p^{-1}$ |
| $\mu$ | Recovery rate | (7 days)<sup>-1</sup> | <sup>5</sup> |
| $\beta_I$ | transmissibility rescaling according to the infectious stage | 0.51 for $I_{p,sc}$, $I_{sc}$<br>1 for $I_{p,c}$, $I_c$ | <sup>6</sup> |
| $\omega_s$ | Transmission risk by setting (network layer) | 1 for $H$ layer<br>0.3 for $C$ layer<br>0.5 otherwise | <sup>1</sup> |

**Table S2.** Vaccine effectiveness parameters and their baseline values.

| Parameter | Description | Baseline value | Source |
| --- | --- | --- | --- |
|  |  | (other explored values) |  |
| $r_{s,1}$ | reduction in susceptibility in the partial-protection stage | 0.52 (0.35, 0.7) | from $VE_{s,1}=48\%$, in the middle of the range of estimates after one dose in <sup>7</sup> (from 30%, 65%, worst and best estimates from <sup>7</sup>, respectively) |
| $\tau_0$ | Average duration of the no-protection stage after first-dose inoculation | $(2 \text{ weeks})^{-1} ((1 \text{ week})^{-1})$ | |
| $r_{s,2}$ | reduction in susceptibility in the maximum-protection stage | 0.3 (0.2, 0.47) | from $VE_{s,2}=70\%$, in the middle of the range of estimates after two doses in <sup>7</sup> (from 53%, 80%, worst and best estimates from <sup>7</sup>, respectively) |
| $r_{c,2}$ | reduction in the probability of developing clinical symptoms in the maximum-protection stage | 0.9 (0.4, 0.95) | from $VE_{sp,2}=73\%$, in the middle of the range of estimates after two doses in <sup>7</sup> (from 60%, 95%, worst and best estimates from <sup>7</sup>, respectively) |
| $\tau_1$ | Average duration of the intermediate-protection stage | $(3 \text{ weeks})^{-1} ((4 \text{ week})^{-1}, (8 \text{ week})^{-1})$ | <sup>8</sup> |
| $p_v$ | Probability of transition between $S^{v,1}$ and $E^v$ | 0.97 (0.65, 0.82) | Computed from the parameters above and assuming $VE_{sp,1}=53\%$, in the middle of the range of estimates after one dose in <sup>7</sup> (from 35%, 75%, worst and best estimates from <sup>7</sup>, respectively) (*) |
| $r_I$ | Reduction in infection duration | 25% (0) | <sup>9</sup> |

(\*) Mid range estimate in <sup>7</sup>, $VE_{sp}=55\%$ , leads to a value of $p_v > 1$ , thus $VE_{sp}=53\%$ is taken.

#### Test-trace-isolate

We model case detection and isolation, combined with tracing and isolation of contacts according to the following rules:

- As an individual shows clinical symptoms, s/he is detected with probability *p_d,c_*. If detected, case confirmation and isolation occur with rate *r_d_* upon symptoms onset.
- Subclinical individuals are also detected with probability *p_d,sc_*, and rate *r_d_*.
- The index case’s household contacts are isolated, with probability *p_ct, HH_*, the same time the index case is detected and isolated. We assume that these contacts are tested at the time of isolation and among those all subclinical, clinical, pre-subclinical, and pre-clinical cases are detected (testing sensitivity 100%).
- Once the index case is detected, contacts of the index case occurring outside the household are traced and isolated with an average delay *r*^−1^. We define an acquaintance as a contact occurring frequently, i.e. with a frequency of activation higher than *f _a_*. We assume that an acquaintance is detected and isolated with a probability *p_ct, A_*, while other contacts (i.e. sporadic contacts) are detected and isolated with probability *p_ct, sp_*, with *p_ct, A_* > *p_ct, sp_*. We assume that traced contacts are tested at the time of isolation and among those all subclinical, clinical, pre-subclinical, and pre-clinical cases are detected (testing sensitivity 100%).
- Only contacts (among contacts occurring both in household and outside) occurring within a window of *D* days before index case detection are considered for contact tracing.
- The index-case and the contacts are isolated for a duration *d _I_* (for all infected compartments) and *d*_¿_ (for susceptible and recovered compartments). Contacts with no clinical symptoms have a daily probability *p_drop_* to drop out from isolation.
- For both the case and the contacts, isolation is implemented by assuming no contacts outside the household and transmission risk per contact within a household reduced by a factor *ι*.

Parameter values are reported in Table S3 and Table S4 for baseline and enhanced TTI, respectively.

**Table S3.** Model for test, trace, isolation. Parameters and their values for the baseline case.

| Parameter | Description | Value | Source |
| --- | --- | --- | --- |
| $p_{d,c}$ | Probability that a clinical case is detected | 0.5 | |
| $p_{d,sc}$ | Probability that a subclinical case is detected | 0.1 | |
| $r_d$ | For detected cases, rate of detection, confirmation and beginning of isolation | $0.28 = (3.6 \text{ days})^{-1}$ | Average time from onset to testing is 2.6 days <sup>10</sup>. We assume one day to have the test results. |
| $D$ | Length of the period preceding index-case confirmation, used to define a contact | 6 days (*) | $\simeq 2 \text{ days} + \frac{1}{r_d}$ (a person is considered to be contact if s/he entered in contact with the index case during a window of 2 days preceding symptoms onset) |
| $p_{ct,HH}$ | Probability that household contacts of an index case are identified and decide to isolate | 0.7 | |
| $p_{ct,A}$ | Probability that acquaintances of an index case are identified and decide to isolate | 0.08 | Calibrated to get $\simeq 2.8$ contacts per index case on average<br>(assumed $p_{ct,Oth} = p_{ct,A}/10$) |
| $p_{ct,Oth}$ | Probability that sporadic contacts of an index case are identified and decide to isolate | 0.008 | Calibrated to get $\simeq 2.8$ contacts per index case on average<br>(assumed $p_{ct,Oth} = p_{ct,A}/10$) |
| $r_{ct}$ | Rate of detection and isolation of contacts outside household | $0.9 = (1.1 \text{ days})^{-1}$ | |
| $f_a$ | Threshold frequency to define a contact as an acquaintance | 1/7 days | |
| $p_{do}$ | Probability to drop out from isolation for individuals that are not clinical | 0.13 | 11 |
| $\iota$ | Reduction in household transmission during isolation | 0.5 | |
| $d_i, d_I$ | Duration of isolation for an individual that is not infectious, duration of isolation for an individual that is infectious | 10 days, 10 days | 12 |

**Table S4.** Model for case test, trace, isolation. Parameters and their values for the case of enhanced TTI. Only values different from the baseline case are reported.

| Parameter | Description | Baseline value (other explored values) |
| --- | --- | --- |
| $p_{d,c}$ | Probability that a clinical case is detected | 0.7 (1) |
| $p_{d,sc}$ | Probability that a subclinical case is detected | 0.3 (0.5) |
| $r_d$ | For detected cases, rate of detection, confirmation and beginning of isolation | $0.9 = (1.1 \text{ days})^{-1}$ |
| $p_{ct,HH}$ | Probability that household contacts of an index case are identified and decide to isolate | 1 |
| $p_{ct,A}$ | Probability that acquaintances of an index case are identified and decide to isolate | 0.24 |
| $p_{ct,Oth}$ | Probability that sporadic contacts of an index case are identified and decide to isolate | 0.024 |
| $p_{do}$ | Probability to drop out from isolation for individuals that are not clinical | 0 |

#### Vaccination strategies

We provide here in the following the parameters values for the vaccination strategies detailed in the Methods section of the main paper.

**Table S5.**
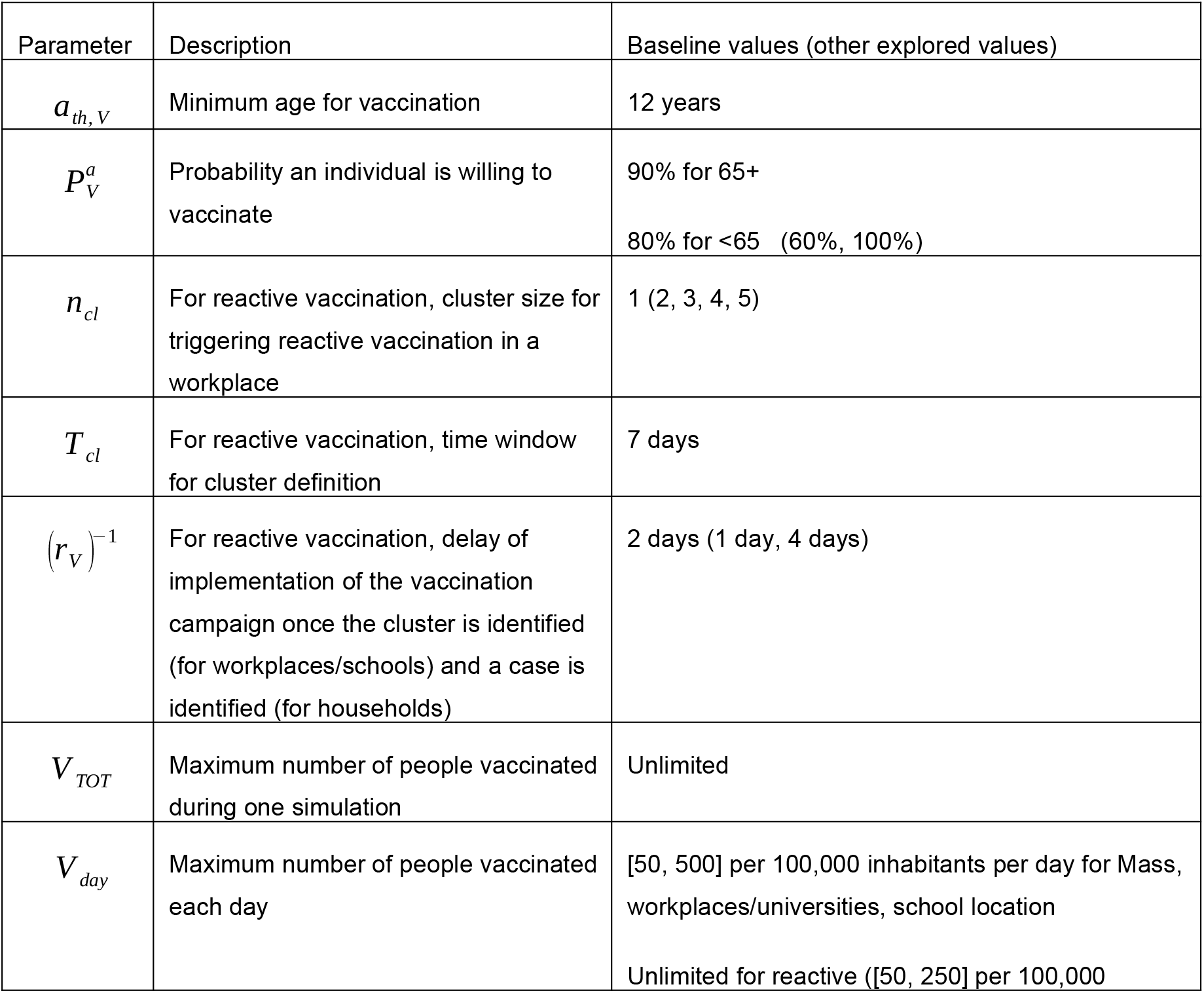
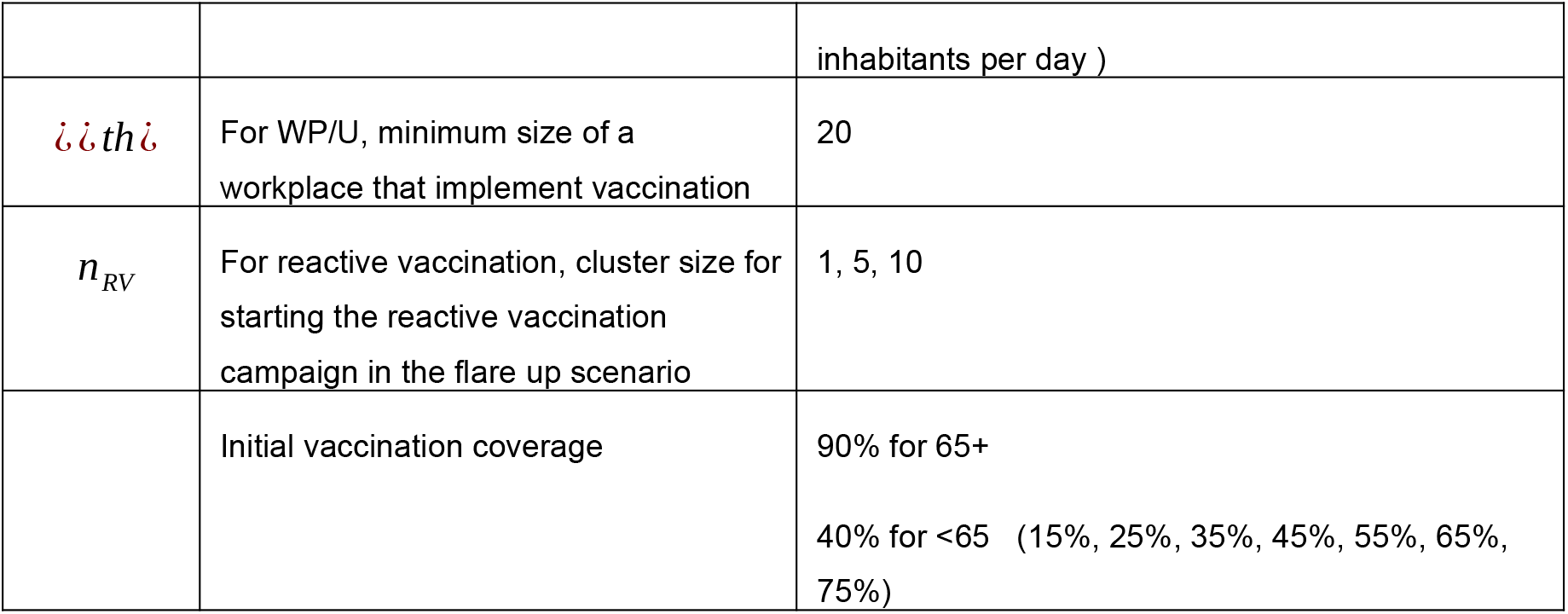
Vaccine administering. Parameters and their baseline values.

| Parameter | Description | Baseline values (other explored values) |
| --- | --- | --- |
| $a_{th,V}$ | Minimum age for vaccination | 12 years |
| $p_v^a$ | Probability an individual is willing to vaccinate | 90% for 65+<br>80% for <65 (60%, 100%) |
| $n_{cl}$ | For reactive vaccination, cluster size for triggering reactive vaccination in a workplace | 1 (2, 3, 4, 5) |
| $T_{cl}$ | For reactive vaccination, time window for cluster definition | 7 days |
| $(r_v)^{-1}$ | For reactive vaccination, delay of implementation of the vaccination campaign once the cluster is identified (for workplaces/schools) and a case is identified (for households) | 2 days (1 day, 4 days) |
| $V_{TOT}$ | Maximum number of people vaccinated during one simulation | Unlimited |
| $V_{day}$ | Maximum number of people vaccinated each day | [50, 500] per 100,000 inhabitants per day for Mass, workplaces/universities, school location<br>Unlimited for reactive ([50, 250] per 100,000) |
|  |  | inhabitants per day ) |
| $\mathbb{I} \mathbb{I} th \mathbb{I}$ | For WP/U, minimum size of a workplace that implement vaccination | 20 |
| $n_{RV}$ | For reactive vaccination, cluster size for starting the reactive vaccination campaign in the flare up scenario | 1, 5, 10 |
|  | Initial vaccination coverage | 90% for 65+<br>40% for <65 (15%, 25%, 35%, 45%, 55%, 65%, 75%) |

### Details on the epidemic simulations

A schematic representation of the main program and of the simulation code and of the algorithm used for a single stochastic realisation are shown in Figure S1 and S2, respectively. Simulations are discrete-time and stochastic. At each time step, corresponding to one day, three processes occur (Figure S2):

*Vaccination Step:* vaccines are administered according to the strategy or the strategies’ combination.

*Testing Step*: cases are detected and isolated; contacts (within and outside household) are identified and isolated; isolated individuals get out from isolation.

*Transmission Step*: infectious status of nodes is updated. This includes transmission, recovery and transition through the different stages of the infection (e.g. from exposed to pre- symptomatic, from pre-symptomatic to symptomatic).

**Figure S1.**
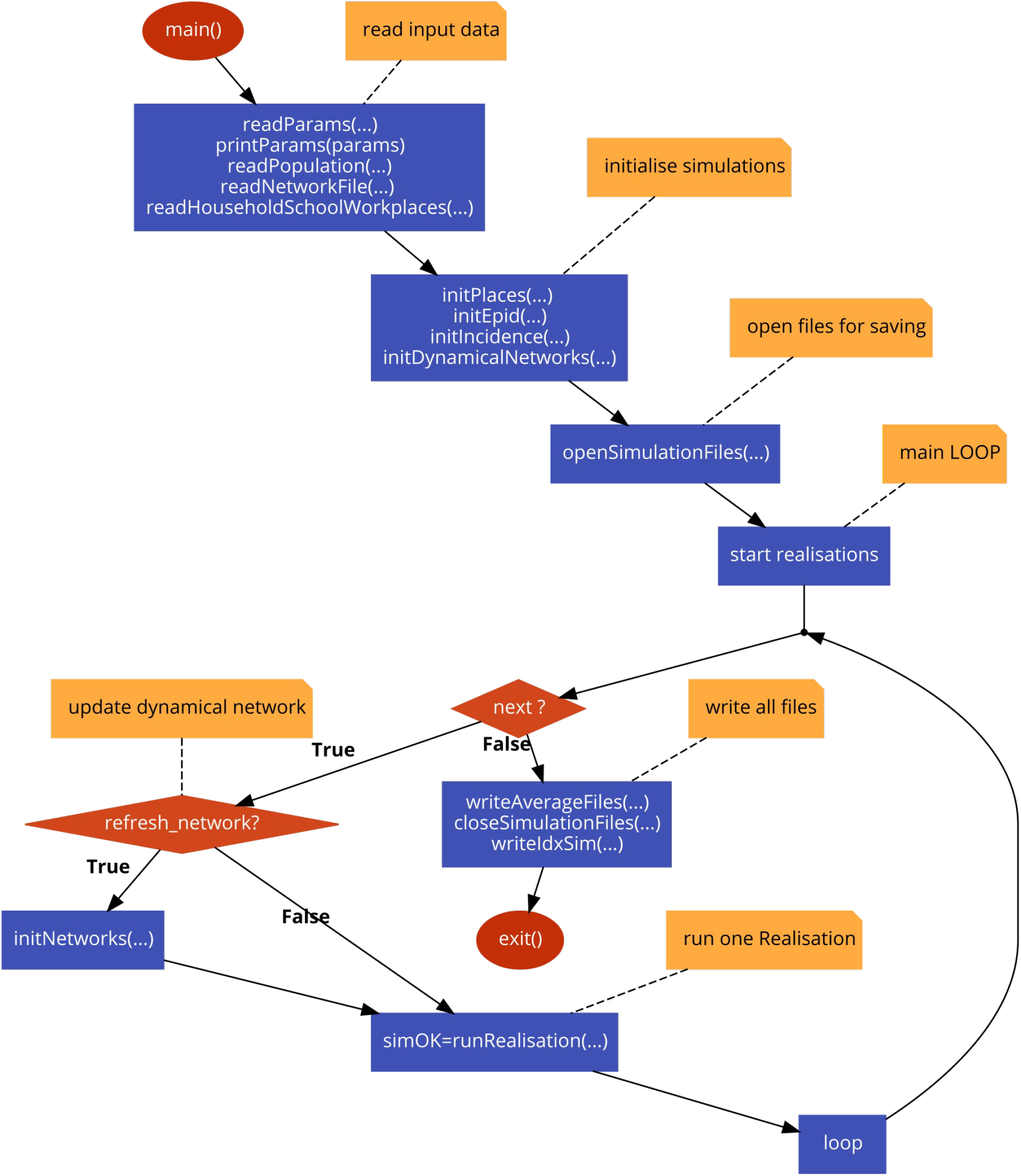
Algorithm of the main program. (drawn with code2flow.com)

**Figure S2.**
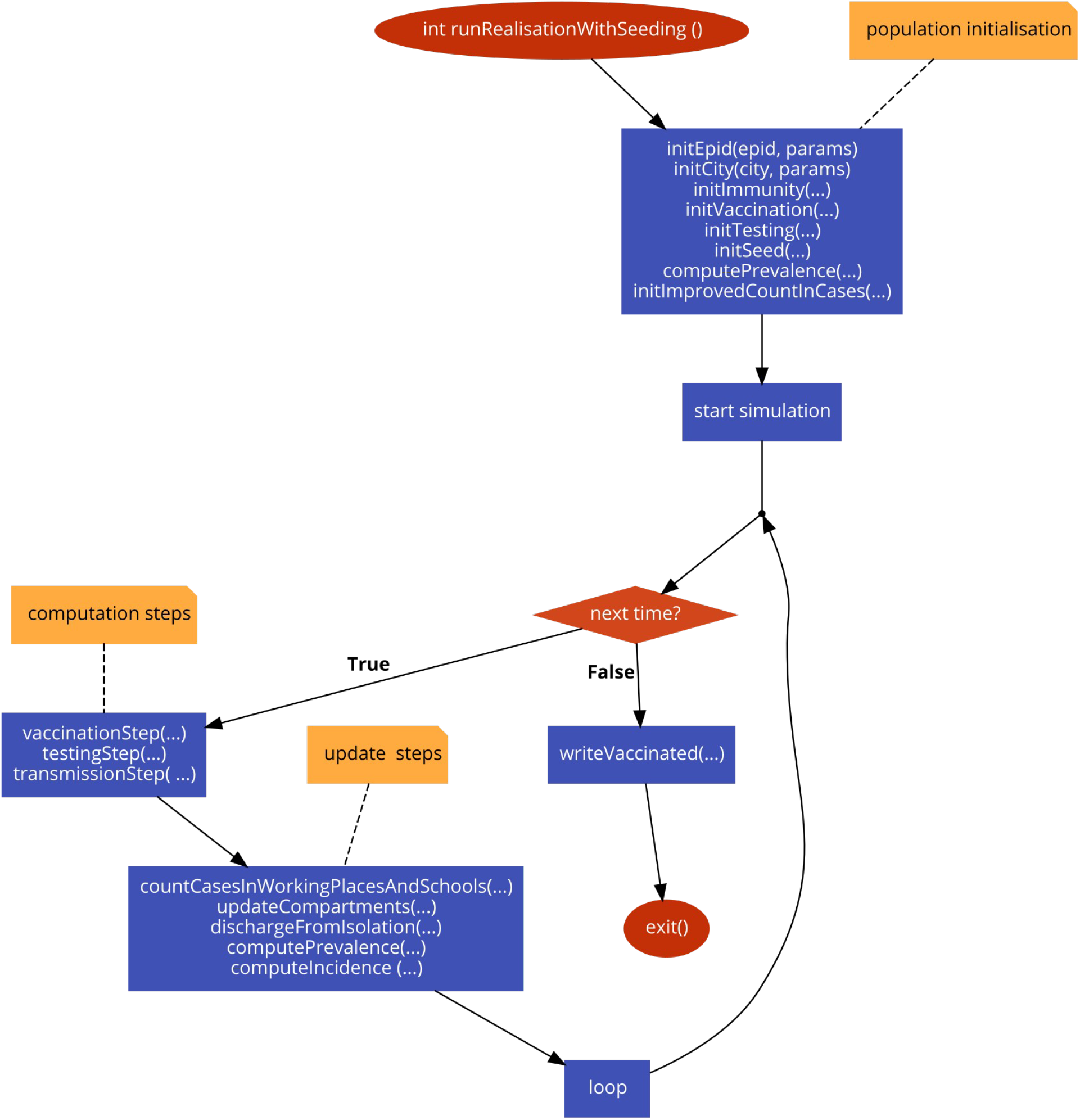
Algorithm for one stochastic realisation. (drawn with code2flow.com)

A single-run simulation is executed with no modelled intervention, until the desired immunity level is reached. This guarantees that immune individuals are realistically clustered on the network. We added some noise, by reshuffling the immune/susceptible status of 30% of the nodes to account for travelling, infection reintroduction from other locations and large gathering with consequent super-spreading not accounted for by the model. In Figure 2 and 3 of the main paper, all processes (transmission, TTI, vaccination) are simulated from the beginning of the simulation. In the scenario with virus re-introduction (Figure 4), TTI and mass vaccination are modelled from the beginning. TTI is enhanced from the detection of the first case. Reactive vaccination starts after the detection of the first *n_RV_* cases, with *n_RV_* =1,5,10 explored.

We vary COVID-19 transmission potential by tuning the daily transmission rate per contact *β*. The reproductive number *R* is computed numerically as the average number of infections each infected individual generates throughout its infectious period at the beginning of the simulation. We tune *β* to have the desired *R* value (1.6 for the analysis in the main text) for the reference scenario - i.e. with only vaccination at the start. We then compare different vaccination strategies at the same value of transmissibility *β*.

To calibrate the duration of the pre-symptomatic stage from ^2^ (value reported in Table S1) we generated an output file containing the list of all transmission events with the infection status of the infector. The proportion of pre-symptomatic transmission was computed as the fraction of transmission events with infector in the compartment *I _p,sc_* or *I _p,c_* over all infection events.

## Additional Results

### Comparison between reactive and non-reactive vaccination strategies Vaccinated settings

We provide here a detailed analysis of the time evolution of the number of settings where vaccines are deployed in the context of reactive vaccination.

**Figure S3.**
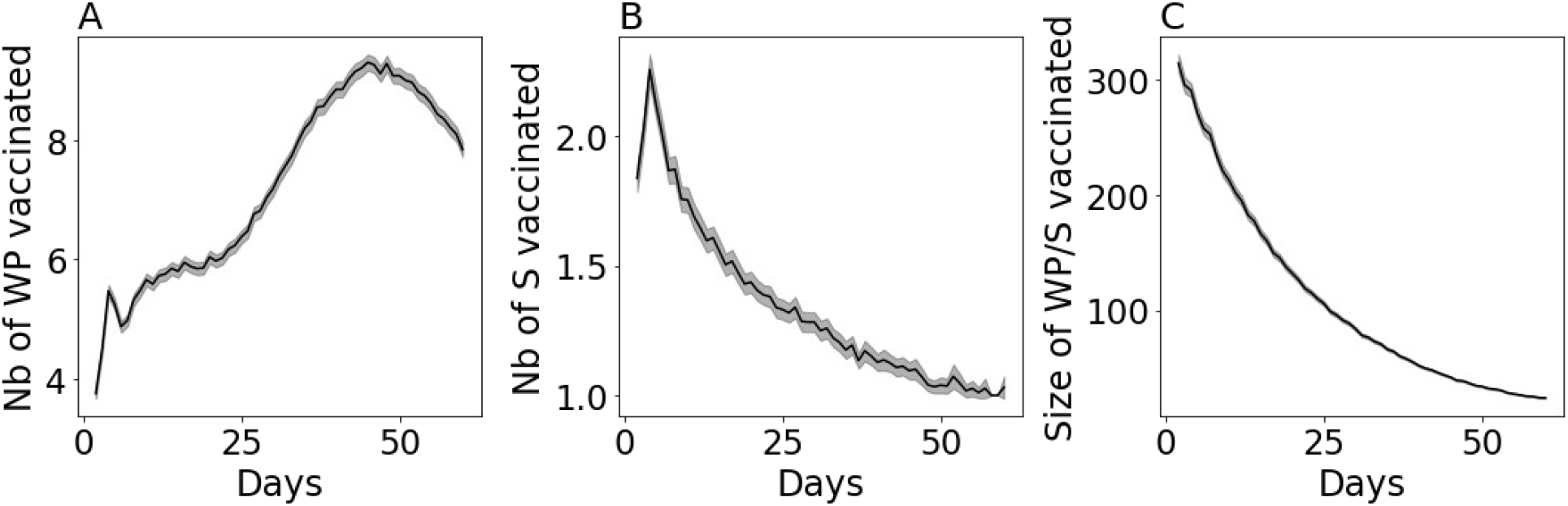
**A, B** Daily number of workplaces (A) and schools (B) where vaccines are deployed in the context of reactive vaccination. **C** Size of workplaces/Schools where vaccines are deployed as a function of time. Values are means over 2000 stochastic simulations. Shaded areas indicate the standard errors.

### Sensitivity analysis

We compare here reactive vaccination with non-reactive vaccination strategies under a variety of epidemic scenarios.

In Figure S4 we compare all strategie at equal number of doses over the two month period, exploring the impact of the following parameters: reproductive ratio, immunity level of the population, repartition of contacts across settings due to social distancing, incubation period, effect of the vaccine in the infection duration, vaccine uptake, and time between doses.

Except when otherwise indicated, parameters are the ones of Figure 2 D, E of the main paper, i.e. intermediate vaccination coverage at the beginning (∼45% of the whole population vaccinated), moderate/high initial incidence and the vaccine uptake to be the same for all strategies. Increasing the transmissibility or initial immunity reduces the impact of reactive vaccination (panel A, B). In panel C we explore the impact of teleworking and reduction in community contact by comparing the baseline scenario with scenarios with no or stronger restrictions. Reactive vaccination becomes more effective when no restriction is in place - although the effect is not strong. This is likely due to the enhanced role of workplaces as a setting of transmission when no teleworking is in place, thus bringing to an increased benefit of reactive vaccination targeting this setting. In panel D, we analyse the impact of the choice of the incubation period exploring incubation period values ranging from 5.1 ^3^ up to 6.3 ^4^. We find that results are highly robust to the choice of the parameters, within this range. In panel E we compare different hypotheses regarding the effect of the vaccine on the infection duration, i.e. the baseline case with the case in which the vaccine induces no reduction in the infectious duration. Also in this case, results are robust. Eventually in panel F we compare different levels of vaccine uptake, assuming uptake to be the same in all strategies as in Figure 2 of the main paper. The impact of vaccination increases with the uptake, the effect being stronger for the reactive strategy. Eventually, we compare in panel G different timing for vaccine-induced immunity to become effective. Specifically, we consider a case in which partial protection against infection mounts one week after the first dose. Assuming an incubation period of ∼6 days, this would be consistent with no reduction in cases observed ∼2 weeks after first-dose inoculation, as reported by some real vaccine effectiveness studies ^14, 15^. We then consider a longer interval between doses (i.e. 4 and 8 weeks). Assuming that protection against infection starts one week after first-dose inoculation leads to a higher impact of vaccination for all four vaccination strategies.

**Figure S4.**
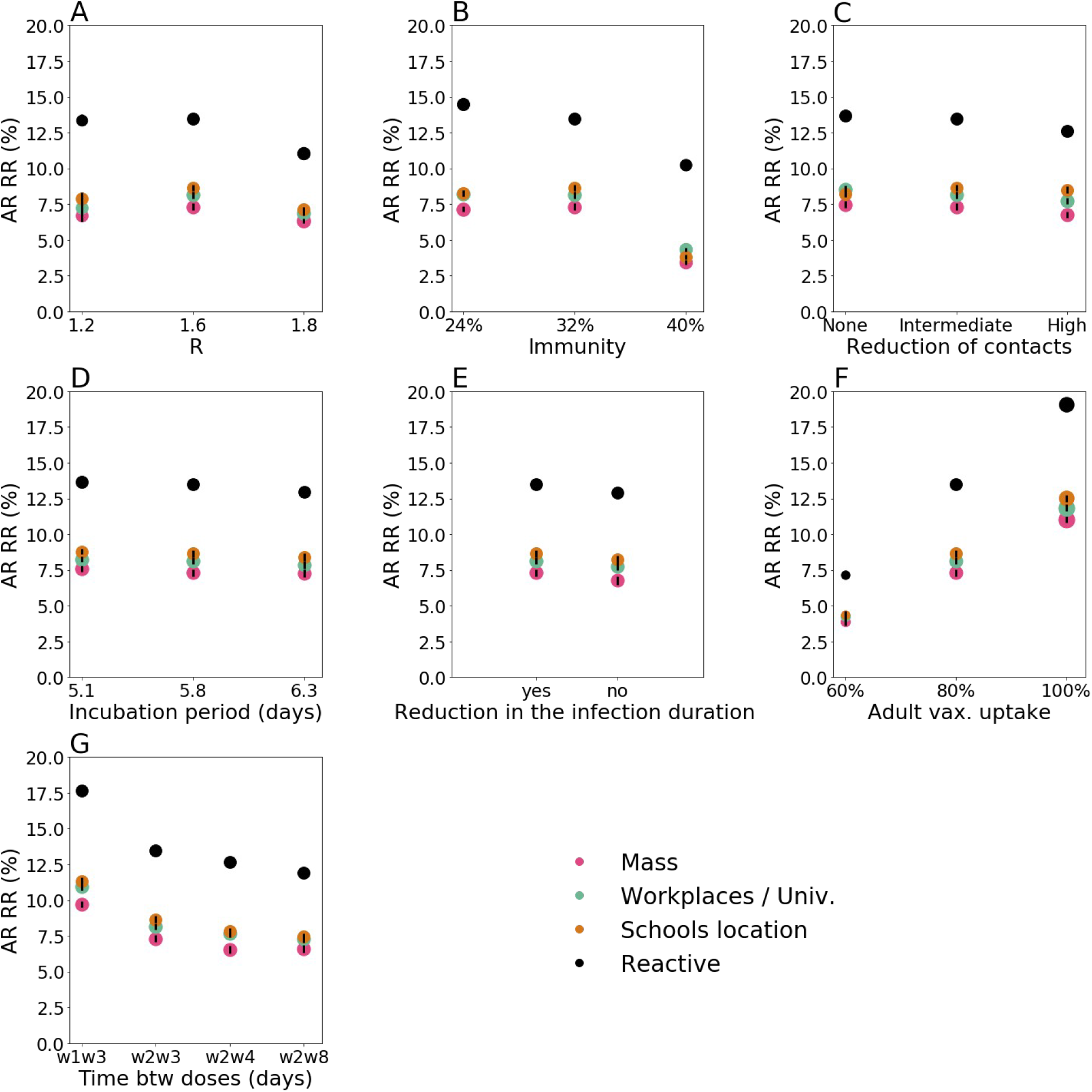
Relative reduction (RR) in the attack rate (AR) after two months for all strategies at equal number of doses. RR is computed with respect to the reference scenario with initial vaccination only. Different parameter values and modelling assumptions are compared. Vaccination rate for mass, workplaces/universities and school location vaccination is set to the average value recovered for reactive vaccination. Exceptions made for the parameter explored in each panel - indicated in the x-axis -, all parameters are as in panel D, E of Figure 2 of the main paper. Parameters explored are: **A** reproductive ratio; **B** natural immunity of the population at the start; **C** repartition of contacts across settings due to social distancing (*Intermediate* is the baseline scenario, while *high* is given by a 30% reduction in community contacts and a 20% of individuals doing teleworking); **D** incubation period, **E** vaccine-induced reduction in infection duration (yes, no), **F** vaccine uptake, and **G** time between doses - the x-axis labels wNwM indicates the average number of weeks, N and M, of no protection following first dose inoculation and of intermediate vaccine effectiveness, respectively. Values are means over 2000 stochastic simulations. Error bars are derived from the standard errors of the AR. These are smaller than the size of the dots in almost all cases.

The impact of reactive vaccination and its demand in terms of vaccine doses varies depending on the incidence level. In Figure S5 A-C we compare all strategies under a scenario of flare-up of cases. We assumed three cases were infected at the beginning. Panels A, B of Figure S5 are the analogous of panel B, E of the Figure 2 of the main paper and are obtained with the same parameters exception made for the value of initial incidence - notably, we assumed intermediate vaccination coverage at the beginning (∼45% of the whole population vaccinated) and the vaccine uptake to be the same for all strategies. Panel A shows the relative reduction in the attack rate after two months as a function of the number of first daily doses, while Panel B compares the incidence profiles under different strategies at equal number of vaccine doses. The relative reduction produced by reactive vaccination is close to the one produced by mass, school location and workplaces/universities. Panel C shows the number of vaccines deployed each day for reactive vaccination and the number of workplaces/schools where vaccines are deployed. These are initially low and increase gradually with incidence.

We then explore the impact of the reactive vaccination within the flare-up case in varying the different parameters. Specifically, we compare all strategies at an equal number of doses, varying the level of social distancing (Figure S5 D, analogously to Figure S4 C) and the timing for the immunity to mount after the first-dose vaccination (Figure S5 E, analogous to Figure S4 G). For certain parameter values reactive vaccination produces a higher relative reduction in the attack rate compared with non-reactive strategies. This is the case for instance when the development of vaccine immunity is rapid, and when no social restrictions are in place. In other cases it produces comparable effect. This is the case for instance of long delays between doses. Finally, in Figure S5 F, we provide an overview of the impact of initial incidence. As initial incidence increases the advantage of the reactive vaccination compared with the non-reactive strategies increases.

**Figure S5.**
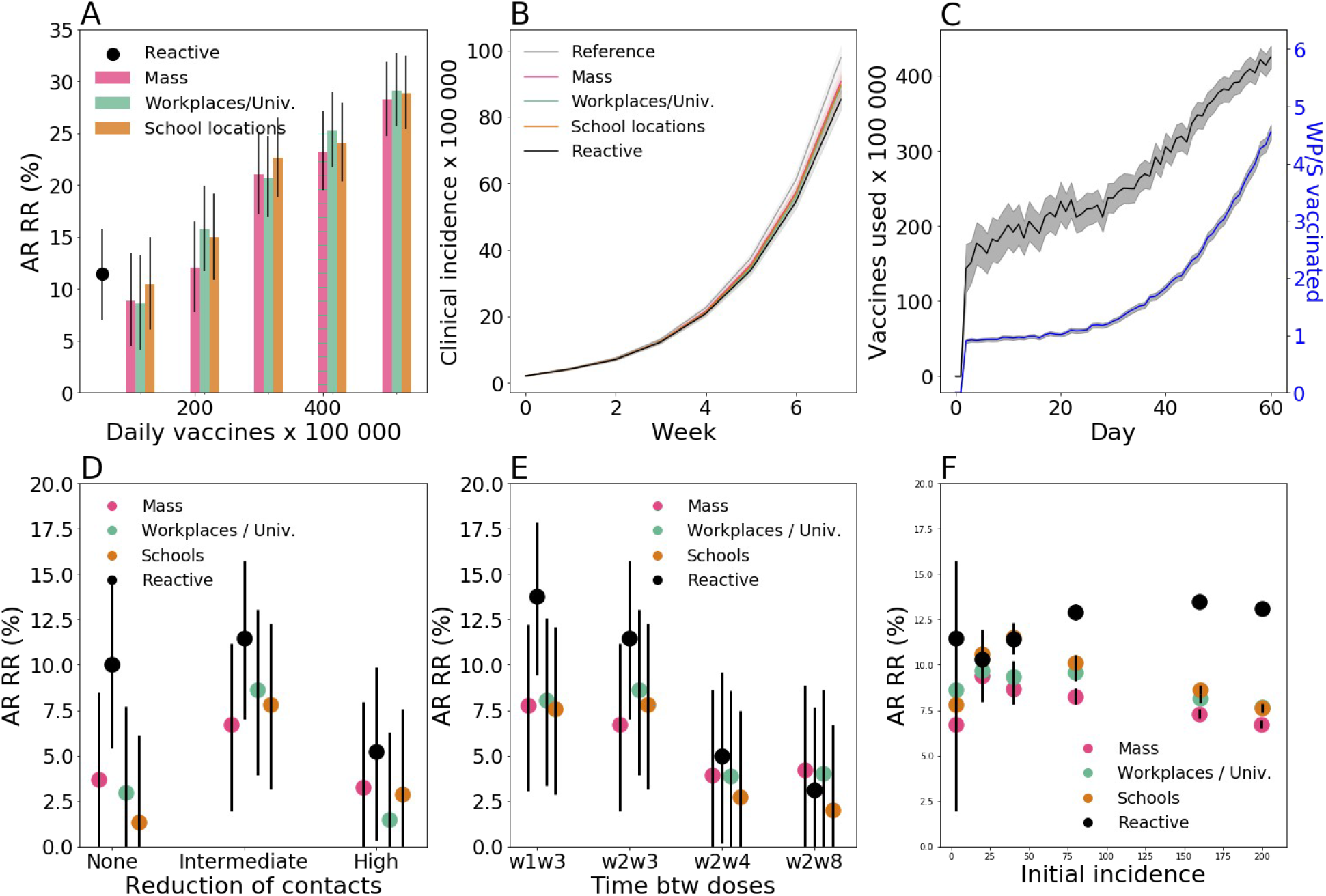
**A** Relative reduction (RR) in the attack rate (AR) over the first two months for all strategies as a function of the vaccination pace. RR is computed with respect to the reference scenario, with initial vaccination only. AR is computed from clinical cases. **B** Incidence of clinical cases with different vaccination strategies. The reactive and non-reactive scenarios plotted are obtained with the same average daily number of vaccines, i.e. 55 daily first-dose vaccinations per 100,000 inhabitants. **C** Number of daily first-dose vaccinations, and workplaces/schools (WP/S in the plot) where vaccines are deployed. **D** AR RR after two months according to the repartition of contacts across settings due to social distancing - *Intermediate* is the baseline scenario, while *high* is given by a 30% reduction in community contacts and a 20% of individuals doing teleworking. **E** AR RR after two months according to the timing for the immunity to mount after first-dose vaccination - the x-axis labels wNwM indicates the average number of weeks, N and M, of no protection following first dose inoculation and of intermediate vaccine effectiveness, respectively. **F** AR RR after two months according to the initial incidence for all strategies at equal number of doses. In panels D-F all strategies are compared at equal number of doses. All panels, except for panel F, consider a flare up scenario, where the epidemic is seeded with 3 infectious individuals. All other parameters are as in panels D, E of Figure 2 of the main paper. Values are means over 2000 stochastic simulations. Error bars in panels A, D, E, F are derived from the standard errors of the AR. Shaded areas in panels B, C, indicate the standard errors.

### Additional epidemic outcomes

Based on the estimated incidence of clinical cases per day provided by the transmission model, we infer outcomes related to hospital, namely hospital and intensive care unit (ICU) entries, beds in ICU ward, and deaths. We use age-dependent hospital admissions (ICU and non-ICU) risks estimated by ^16, 17^ and ICU admission risks for hospitalised patients based on SI-VIC extract ^18^. Hospital admissions risks were adjusted to apply only to clinical cases ^19^ and to account for vaccine effectiveness for hospitalisation for zero, half (1 dose) and full (2 doses) vaccination. We assumed the vaccine efficacies for hospitalisation were equal to 83% and 94% for half and full vaccinations, respectively^7^. We also assume that the hospital admission risk increases by 80% with the Delta variant compared to Alpha ^20^ and by 40% with the Alpha variant compared to the wild type ^21^. Patients who were hospitalised entered the hospital on average 7 days (sd = 3.9 days – Gamma distribution) after the beginning of the infectious phase ^22^. Those who were admitted in ICU enter this unit with a mean delay of 1.69 days (assuming an exponential distribution) ^18^. To estimate the number of occupied beds, we use age-specific mean durations of stay and their corresponding standard deviations in ICU calculated on all the hospitalised cases in the first 9 months of the French epidemic (March-November 2020)^18^. We assume that the standard deviations of ICU lengths of stay were equal to the corresponding mean and do not consider post-ICU care in the estimation of occupied beds. We estimate the number of deaths using hospital and ICU death risks of hospitalised infected persons ^18^. Deaths are delayed in time using the mean delays and standard deviations from hospital or ICU admission to death ^18^. All lengths of stay are supposed to follow a Gamma distribution. Parameters and their values are summarised in Tables S6 and S7.

We also estimate the number of life years and quality-adjusted life years (QALY) lost for each death using life-tables provided by ‘French National Institute of Statistics and Economic Studies’ (INSEE) for 2012-2016 ^23^ and utility measures of each age-group in France ^24^.

**Table S6.** Risks of hospitalisation according to vaccination status, ICU admission and death in general ward and ICU.

| Age Group | Hospitalisation risk for no-vaccination | Hospitalisation risk for half vaccination (1 dose) | Hospitalisation risk for full vaccination (2 doses) | ICU admission risk | Death risk in general ward | Death risk in ICU |
| --- | --- | --- | --- | --- | --- | --- |
| 0-9 | 0.0246 | 0.0115 | 0.0066 | 0.159 | 0.006 | 0.078 |
| 10-19 | 0.0136 | 0.0063 | 0.0037 | 0.159 | 0.006 | 0.078 |
| 20-29 | 0.0364 | 0.0170 | 0.0098 | 0.159 | 0.006 | 0.078 |
| 30-39 | 0.0443 | 0.0207 | 0.0119 | 0.159 | 0.006 | 0.078 |
| 40-49 | 0.0617 | 0.0288 | 0.0166 | 0.219 | 0.016 | 0.103 |
| 50-59 | 0.1697 | 0.0792 | 0.0457 | 0.270 | 0.030 | 0.175 |
| 60-69 | 0.2360 | 0.1101 | 0.0635 | 0.299 | 0.064 | 0.268 |
| 70-79 | 0.5113 | 0.2386 | 0.1377 | 0.235 | 0.140 | 0.366 |
| 79+ | 0.9496 | 0.4431 | 0.2557 | 0.053 | 0.308 | 0.468 |

**Table S7.** Delays from hospitalisation admission in general ward to death or hospital discharge and delays from ICU admission to ICU discharge or death.

| Age<br>Group | Mean los in general<br>ward (sd) | Mean los in general<br>ward for dying<br>people (sd) | Mean los in ICU<br>(sd) | Mean los in ICU for<br>dying people (sd) |
| --- | --- | --- | --- | --- |
| 0-9 | 6.4 (6.8) | 10.6 (12.3) | 12.7 (12.7) | 22.3 (22.3) |
| 10-19 | 6.4 (6.8) | 10.6 (12.3) | 12.7 (12.7) | 22.3 (22.3) |
| 20-29 | 6.4 (6.8) | 10.6 (12.3) | 12.7 (12.7) | 22.3 (22.3) |
| 30-39 | 6.4 (6.8) | 10.6 (12.3) | 12.7 (12.7) | 22.3 (22.3) |
| 40-49 | 6.4 (6.8) | 10.6 (12.3) | 12.7 (12.7) | 22.3 (22.3) |
| 50-59 | 6.4 (6.8) | 10.6 (12.3) | 12.7 (12.7) | 22.3 (22.3) |
| 60-69 | 9.3 (9.1) | 10.6 (12.3) | 16.7 (16.7) | 20.8 (20.8) |
| 70-79 | 11.7 (11.4) | 10.6 (12.3) | 17.5 (17.5) | 18.9 (18.9) |
| 79+ | 15 (13.8) | 10.6 (12.3) | 13.6 (13.6) | 10.6 (10.6) |

Los: Length of stay. Sd = Standard deviation.

Figure S5 shows the relative reductions in the number of hospitalisations, deaths, ICU entries, life-year lost, quality-adjusted life-year lost and ICU bed occupancy at the peak, comparing each vaccination scenario with the reference scenario - i.e. vaccination only at the start. We consider here the high incidence and intermediate vaccine coverage scenario, analogously to Figure 2 D, E of the main paper. The six indicators show a behaviour similar to incidence.

Overall reduction values are smaller. This is expected, since a large proportion of elderly are already vaccinated at the start, and the compared vaccination strategies target a population that is less at risk of severe infection. Still all indicators show the same qualitative behaviour, with reactive vaccination outperforming the non-reactive vaccination strategies at equal number of first-dose vaccination.

**Figure S6.**
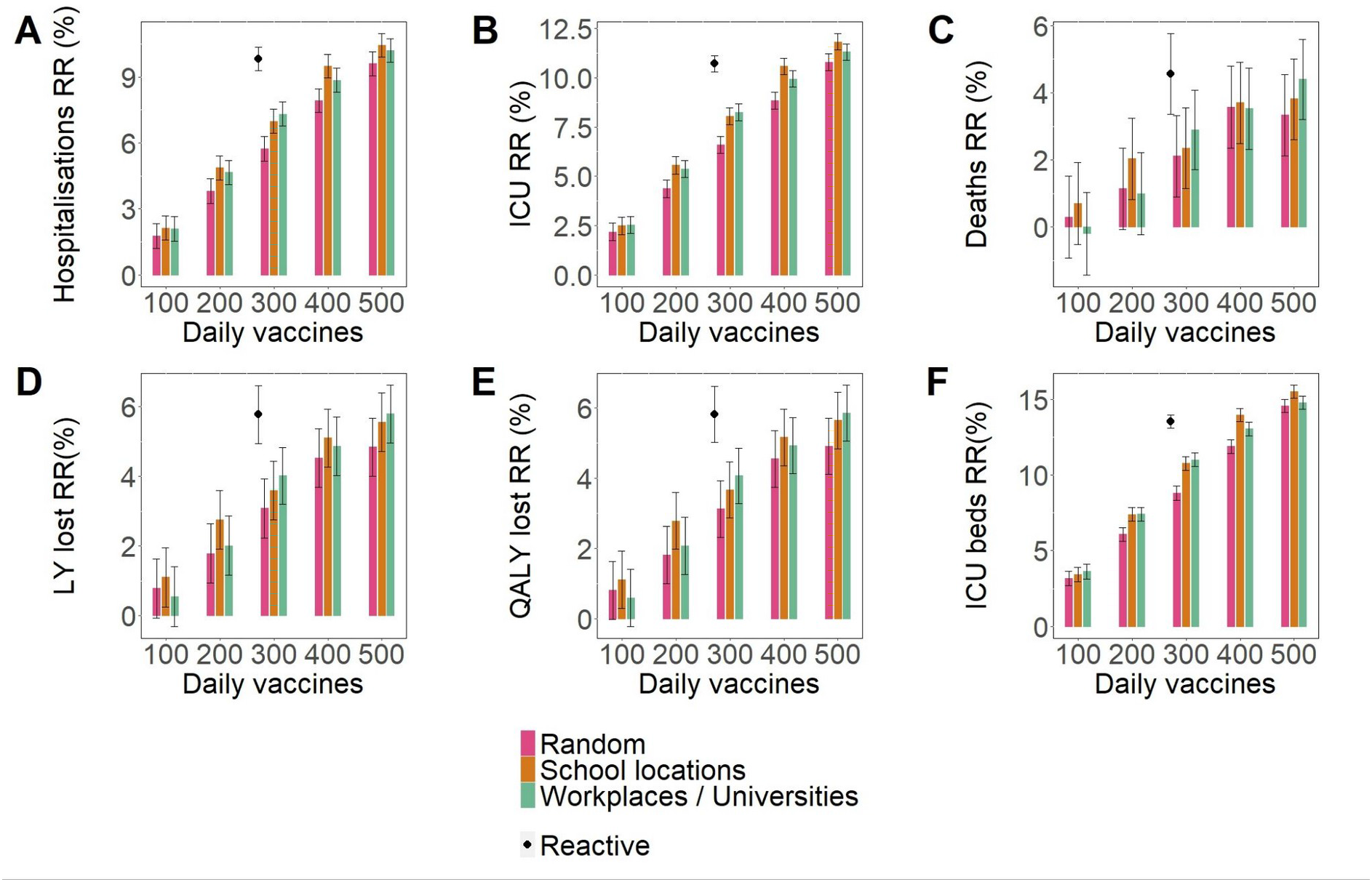
**A, B, C, D, E** Relative reduction (RR) in the cumulative incidence of hospitalisations, intensive care unit (ICU) entries, deaths, life years (LY) lost and quality-adjusted life years (QALY) lost over the first two months for all strategies as a function of the vaccination pace. **F** Relative reduction (RR) in occupied ICU beds at the peak over the first two months for all strategies as a function of the average daily number of first-dose vaccinations for the intermediate vaccination coverage scenario. Values are means over 2000 stochastic simulations. Error bars are derived from the standard errors of the AR. Parameters are the same as in Figure 2 D, E of the main paper.

### Combined reactive and mass vaccination for managing sustained COVID-19 spread

#### Additional results

In Figure S7 we show the incidence curve corresponding to the scenarios analysed in Figure 3 A of the main paper. Mass and combined vaccination with the five different vaccination paces are compared.

**Figure S7.**
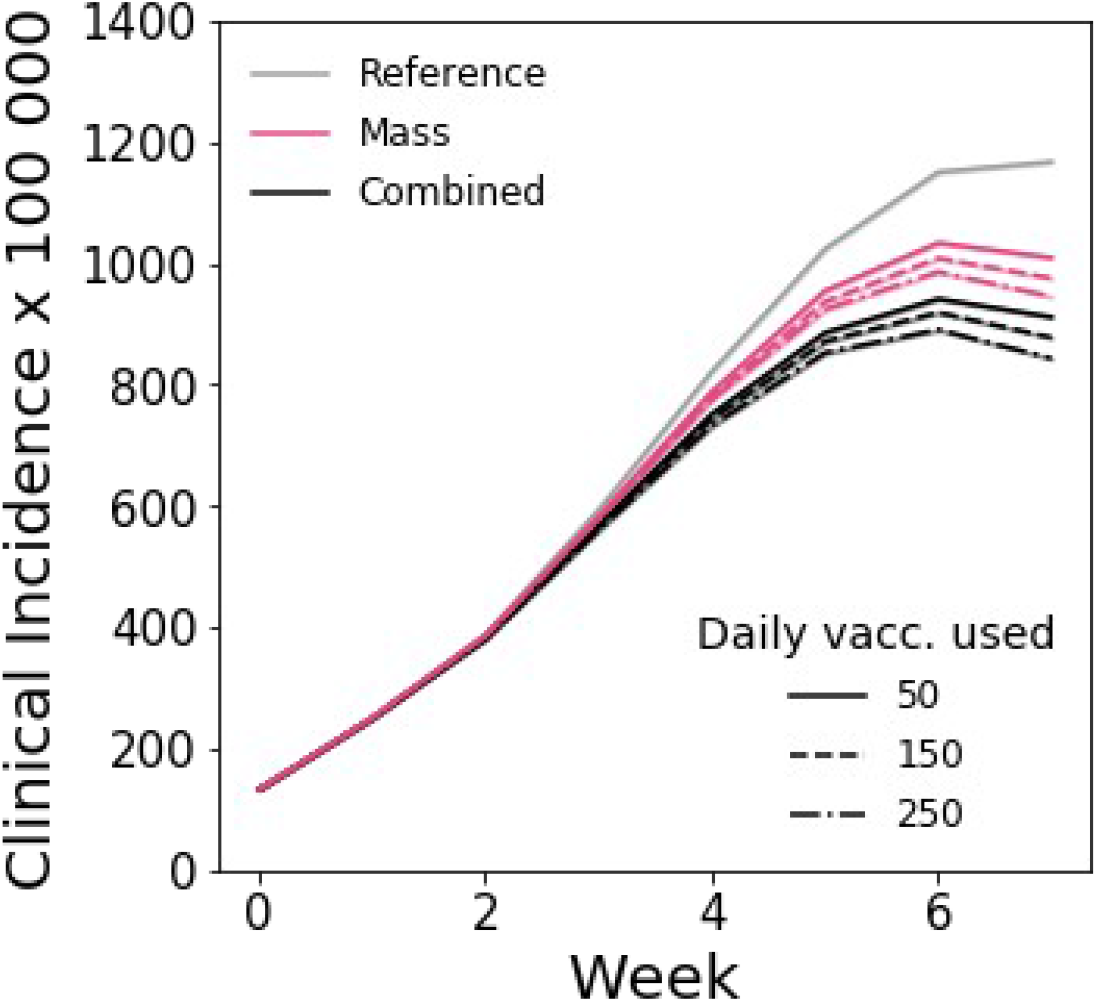
Incidence of clinical cases for mass and combined vaccination strategies for three different vaccination paces. Scenarios are the same as the ones plotted in Figure 3 A. We assumed the following parameters: initial incidence of clinical cases 160 per 100 000 inhabitants; R=1.6; Initial immunity 32%; vaccinated at the beginning 90% and 40% for 60+ and <60, respectively; 10% of individuals are doing teleworking and 5% of community contacts are removed. Values are means over 2000 stochastic simulations. Shaded areas indicate the standard errors - very low for this set of parameters.

### Combined reactive and mass vaccination for managing a COVID-19 flare-up

#### Additional results

In Figure S8 we show the incidence curve corresponding to the scenarios analysed in Figure 4 of the main paper. Mass and combined vaccination with the different vaccination scenarios considered are compared.

**Figure S8.**
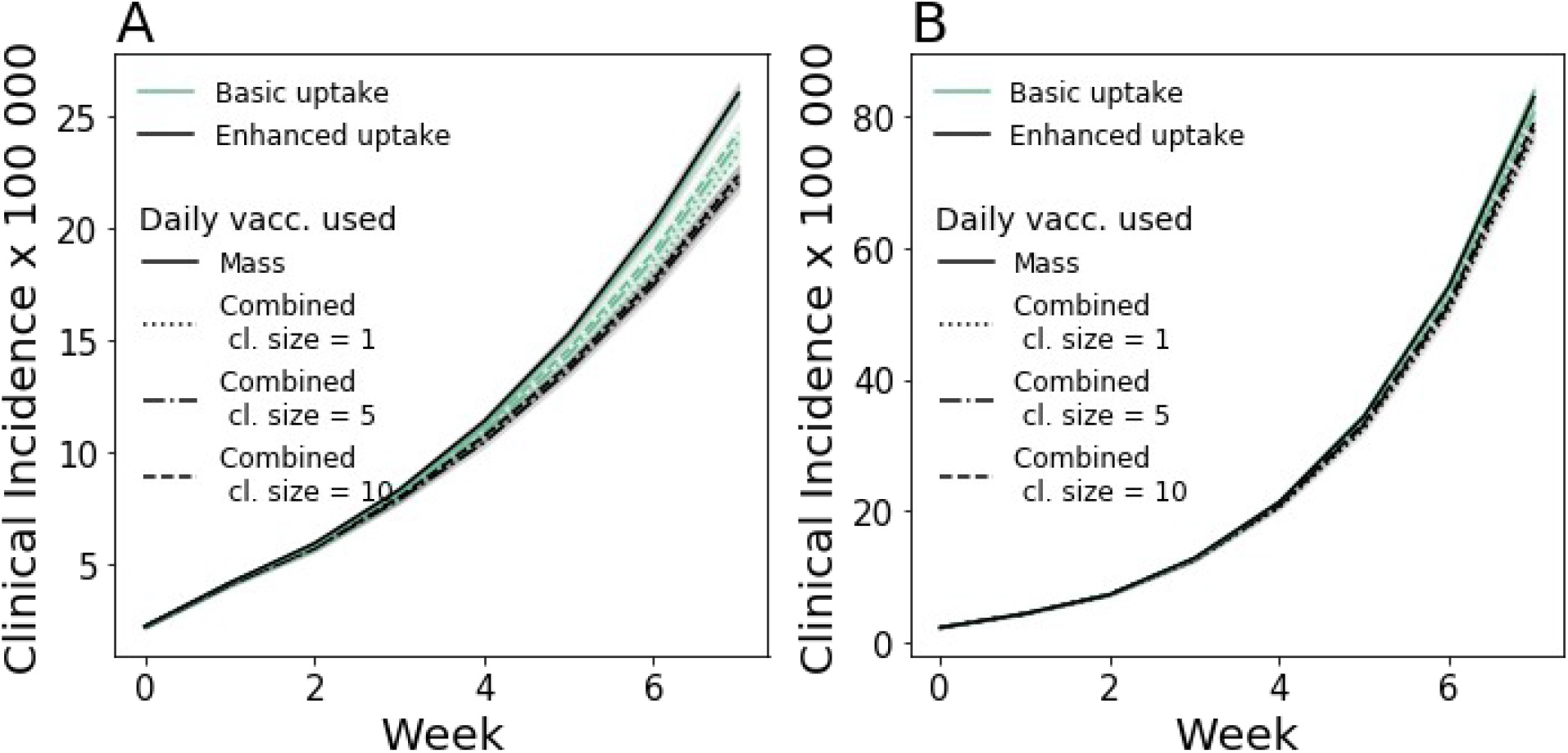
Incidence of clinical cases for mass and combined vaccination strategies for the scenarios analysed in Figure 4 of the main paper. **A** scenario with enhanced TTI. **B** scenario with baseline TTI. In all panels we assumed the following parameters: R=1.6; Initial immunity 32%; 90% for 60+ year old and 40% for <60 years old already vaccinated at the beginning; 10% of individuals are doing teleworking and 5% of community contacts are removed. Values are means over 8000 stochastic simulations. Shaded areas indicate the standard errors.

#### Sensitivity analysis

In Figure S9 we analyse the impact of combined and mass vaccination in a flare up scenario similarly to Figure 4, by varying the hypotheses on virus transmissibility and vaccine escape. Specifically we consider values of the reproductive ratio from 1.2 to 1.8, and both worst and baseline vaccine effectiveness level - the worst vaccine effectiveness level is the same as in Figures 2 I of the main paper. Analogously to Figure 4 we compare the attack rate for combined and mass vaccination, assuming both baseline and enhanced TTI and both baseline and 100% vaccine uptake in the context of reactive vaccination. We consider only the case in which reactive vaccination starts after the detection of the first case. For each set of parameters, scenarios with enhanced TTI and 100% uptake are associated with smaller attack rates and larger difference between mass and combined than the corresponding scenarios with baseline TTI and baseline uptake.

**Figure S9.**
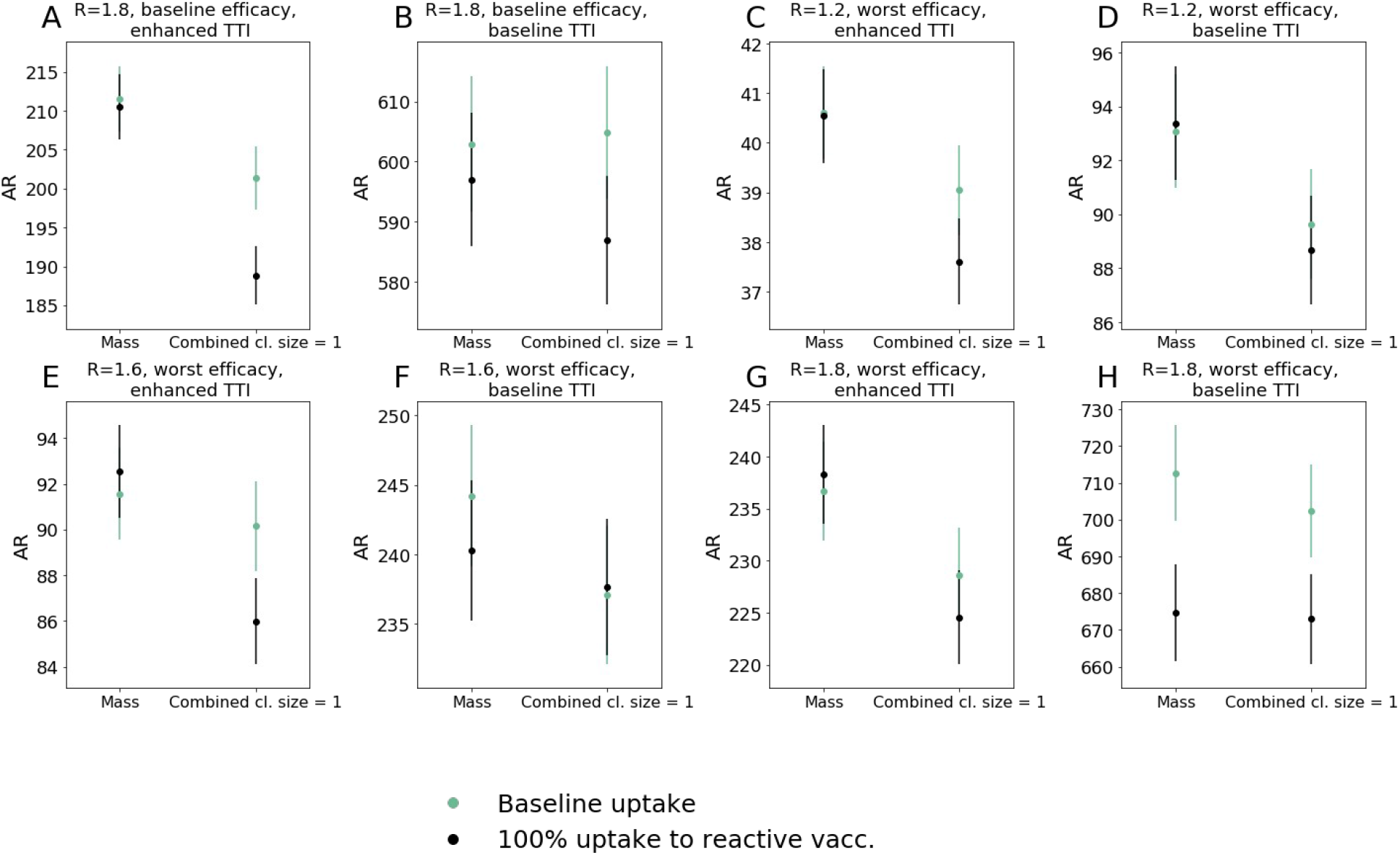
Average attack rate per 100000 inhabitants after two months for the flare up case under different hypotheses: **A**, **B**, R = 1.8 with baseline vaccine effectiveness; **C-H** worst vaccine effectiveness with R = 1.2 (C,D), R = 1.6 (E,F) and R = 1.8 (G,H). The worst vaccine effectiveness scenario is defined as in figure 2 I, i.e. *V E_S_*_,1_=30 %, *V E_SP_*_, 1_=35 %, *V E_S_*_,2_=53 % and *V E_SP_*_, 2_=60 %. We compare enhanced and baseline TTI - A, C, E, G and B, D, F, H, respectively -, as well as baseline and 100% vaccine uptake. In all panels, parameters are the same as in Figure 4: initial immunity 32%; 90% for 60+ year old and 40% for <60 years old already vaccinated at the beginning; 10% of individuals are doing teleworking and 5% of community contacts are removed. Values are means over 8000 stochastic realisations. Error bars are the standard error.

We then investigate the impact of combined and mass vaccination on the extinction of the flare-up. In Figure 10 A we plot the probability of extinction for the scenario considered in Figure 4 A of the main paper (enhanced TTI and 100% vaccine uptake). We find that the probability of extinction is ∼5%, and the difference between mass and combined is ∼0.5%. The probability of extinction progressively increases under more optimistic hypotheses: increase in case detection from 70% and 30% (enhanced TTI) to 100% and 50% (strong TTI) for clinical and subclinical cases, respectively; increase in vaccine effectiveness to the best case scenario considered in Figure 2 I; rapid mounting of the vaccine effect, with partial immunity against infection already present one week after inoculation. In the best-case scenario plotted in panel H, the probability of extinction reaches ∼0.15 and ∼0.18 for mass and combined vaccination, respectively.

**Figure S10.**
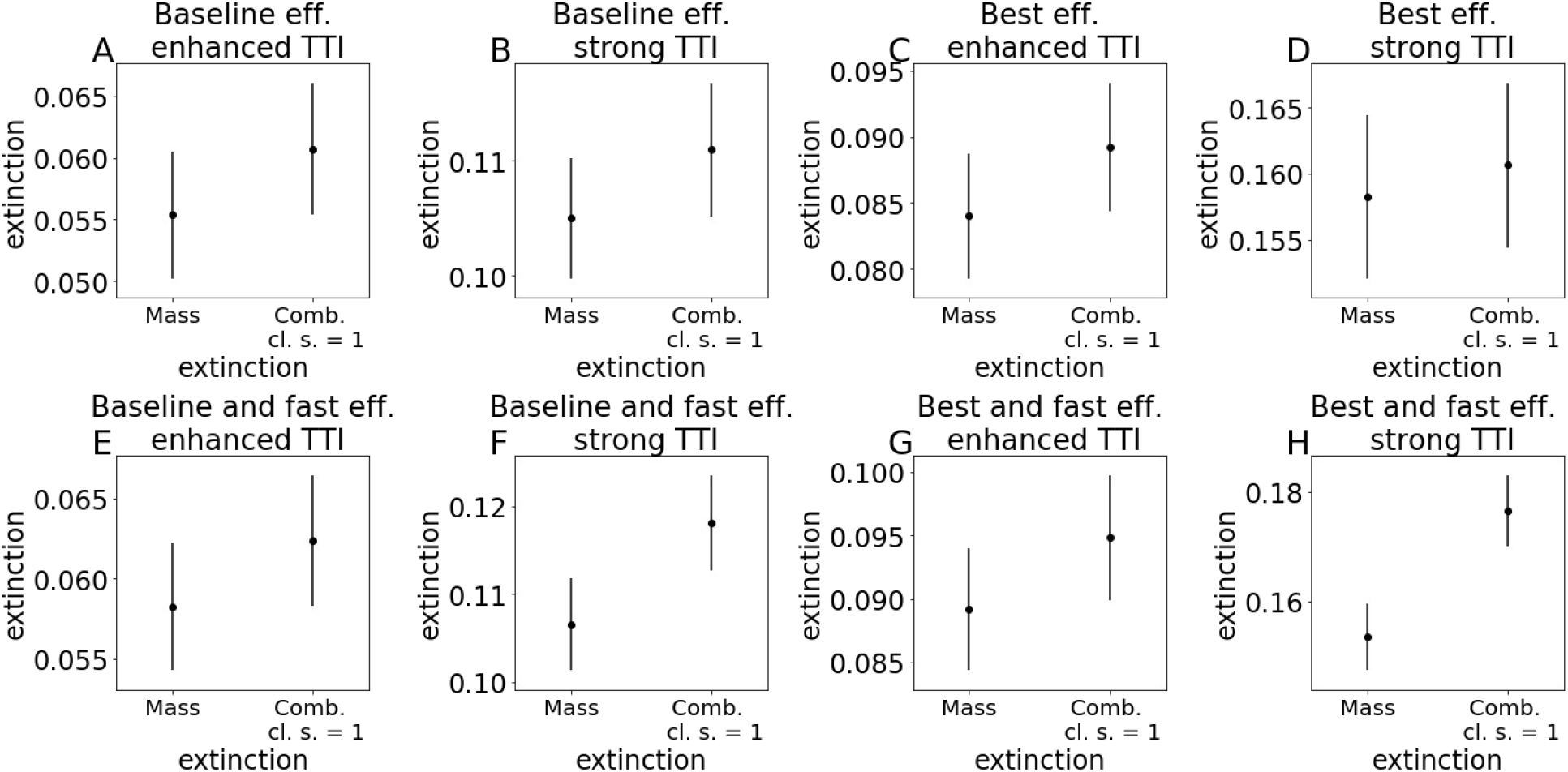
Probability of extinction for an outbreak of three initial cases for mass vs. combined vaccination, compared at an equal number of doses. Three sets of parameters are investigated. 1) Baseline vaccine effectiveness (**A,B,E,F**) vs best effectiveness, i.e. *V E_S_*_,1_=65 %, *V E_SP_*_, 1_=75 %,*V E_S_*_,2_=80 % and *V E_SP_*_, 2_=95 %, (**C,D,G,H);** 2) enhanced TTI (**A,C,E,G**) vs. strong TTI with *p_d,c_*= 1 and *p_d,sc_* = 0.5 (**B,D,F,H**). 3) 2 weeks (baseline) for vaccines to reach partial effectiveness (**A,B,C,D**) vs 1 week (**E,F,G,H**). In all panels, parameters are the same as in Figure 4: initial immunity 32%; 90% for 60+ year old and 40% for <60 years old already vaccinated at the beginning; 10% of individuals are doing teleworking and 5% of community contacts are removed. Values are means over 15000 stochastic realisations. Error bars are the standard error.

